# Inequalities in children’s mental health care: analysis of routinely collected data on prescribing and referrals to secondary care

**DOI:** 10.1101/2022.06.14.22276082

**Authors:** William P Ball, Corri Black, Sharon Gordon, Bārbala Ostrovska, Shantini Paranjothy, Adelene Rasalam, David Ritchie, Helen Rowlands, Magdalena Rzewuska, Elaine Thompson, Katie Wilde, Jessica E Butler

## Abstract

**Background:** One in eight children in the United Kingdom are estimated to have a mental health condition, and many do not receive support or treatment. The COVID-19 pandemic has negatively impacted mental health and disrupted the delivery of care. Prevalence of poor mental health is not evenly distributed across age groups, by sex or socioeconomic groups. Equity in access to mental health care is a policy priority but detailed socio-deomgraphic trends are relatively under-researched.

**Methods:** We analysed records for all mental health prescriptions and referrals to specialist mental health outpatient care between the years of 2015 and 2021 for children aged 2 to 17 years in a single NHS Scotland health board region. We analysed trends in prescribing, referrals, and acceptance to out-patient treatment over time, and measured differences in treatment and service use rates by age, sex, and area deprivation.

**Results:** We identified 18,732 children with 178,657 mental health prescriptions and 21,874 referrals to specialist outpatient care. Prescriptions increased by 59% over the study period. Boys received double the prescriptions of girls and the rate of prescribing in the most deprived areas was double that in the least deprived. Mean age at first mental health prescription was almost 1 year younger in the most deprived areas than in the least. Referrals increased 9% overall. Initially, boys and girls both had an annual referral rate of 2.7 per 1,000, but this fell 6% for boys and rose 25% for girls. Referral rate for the youngest decreased 67% but increased 21% for the oldest. The proportion of rejected referrals increased steeply since 2020 from 17% to 30%. The proportion of referrals accepted for girls rose to 62% and the mean age increased 1.5 years.

**Conclusions:** The large increase in mental health prescribing and changes in referrals to specialist outpatient care aligns with emerging evidence of increasing poor mental health, particularly since the start of the COVID-19 pandemic. The static size of the population accepted for specialist treatment amid greater demand, and the changing demographics of those accepted, indicate clinical prioritisation and unmet need. Persistent inequities in mental health prescribing and referrals require urgent action.

## Background

Childhood and adolescence are key periods in lifetime mental health trajectories (1). Research from the United Kingdom estimates that around 1 in 8 children and young people have a diagnosable mental health condition, and that prevalence has increased over time (2). Half of all long-term mental disorders are thought to start by age 14, and three-quarters by age 24 (1). Despite this, many of these young people do not receive treatment (3,4). The prevalence of mental health conditions is not evenly distributed across the population, with key differences between groups based on age, sex and socioeconomic conditions. Older children, girls and those from deprived areas are all more likely to present with poor mental well-being (5–7). Addressing unequal access to Child & Adolescent Mental Health Services (CAMHS) has been a longstanding Scottish Government priority (8) and mental health and inequalities remain a key concern in the period of transition and recovery from the impacts of COVID-19 (9).

Trends across the UK and around the world suggest increasing prevalence of mental health problems (6) and inequalities in self-reported mental wellbeing by area deprivation (7), although comprehensive and detailed data is lacking. Prescribing of medications used to treat mental health conditions in children has increased in various international contexts(10,11) with notable rises in prescriptions of antidepressants (12,13) and medications to treat Attention Defecit Hyperactivity Disorder particularly among boys (14,15) from the 1990s up until the 2010s. Analysis of mental health prescribing in parts of the United Kingdom has previously been conducted for people of all ages, but this did not include a detailed exploration of trends in younger age groups (16,17). Analysis of psychotropic prescribing rates in young people during 2020 found an initial decrease during the period of national lockdowns, followed by increases particularly for antidepressants (18). The number of young people being treated for mental health complaints has increased over time internationally (19,20) and there is emerging international evidence that this increase has accelerated since the onset of the COVID-19 pandemic (21–23). Aggregated open data on prescribing and referrals to specialist outpatient (Tier 3) services are available in Scotland and confirm a trend of increasing referral through time which has accelerated since the COVID-19 pandemic (24). Previous analysis of one year of specialist outpatient referrals in a single Scottish council region found important demographic patterns, with older children and boys more likely to be referred (25) but did not provide population adjusted rates or a sense of temporal trends which are presented in our analysis. National-level analysis of prescribing and specialist referrals in Scotland has provided high-level summary insights (8,16,26), but analysis of sub-national trends are needed to provide improved local understanding to inform service and workforce planning (27). Here we used individual level data for analysis of a wider range of social and demographic characteristics associated with mental health prescribing and service use.

We describe the rates of prescribing of medications commonly used to treat mental health conditions in children and explore trends in specialist outpatient referrals, rejections, and treatment both before and during the COVID-19 pandemic. We describe social and demographic characteristics for these populations, including sex, age, and home-area deprivation. Finally, we quantify inequality in rates of prescription and referral by area deprivation.

## Methods

### Study Setting

The study setting is the NHS Grampian health board region located in the North East of Scotland which covers the Aberdeen City, Aberdeenshire and Moray local authority areas. The population is roughly evenly split between urban and rural areas and the health board is relatively less deprived than others in Scotland. Some areas are ranked in the 10% most deprived by Scottish Index of Multiple Deprivation (SIMD) (28) but these areas account for just 1% of the health board population, compared with 15% living in areas ranked in the 10% least deprived (29). The annual size of the under-18 aged population in NHS Grampian has remained static at roughly 111,000 individuals throughout this study period (29).

### Data Sources

Prescription data are from the national Prescribing Information System (PIS) (30) which provides records for all medicines which are prescribed or dispensed in a community setting in Scotland which includes most outpatient prescriptions which are commonly dispensed in this setting. Child and Adolescent Mental Health Services (CAMHS) data (31) is an NHS Grampian dataset which is used to report official waiting-list statistics at the national level. Home area-level socioeconomic information is sourced from version 2 of the 2020 SIMD (28) which has been linked to individual records using data zone of residence at the time of prescription or referral. Rates for outcomes have been calculated using official mid-year population estimates for the NHS Grampian health board region (29).

Linkages between datasets were made using the Community Health Index (CHI) number which is a unique identifier from the CHI population register for Scotland (32). Data linkage and management was conducted by Grampian Data Safe Haven (DaSH) (33) staff on a secure server using accredited procedures to collect, link and pseudonymise patient-level records. Access to de-identified records was allowed through the Trusted Research Environment (TRE) for only named and approved researchers and outputs in the form of aggregate summary data, tables or figures were subject to disclosure control procedures to reduce the risk of identifiable information being shared.

### Study Population

Records were available for all mental health prescriptions and referrals to specialist CAMHS between 1^st^ January 2015 and 31^st^ October 2021 for individuals resident in the NHS Grampian Health Board region. This study includes individuals with at least one mental health prescription or specialist CAMHS referral during the study period. Records were restricted to individuals aged between 2 and 17 years. Under 2 year olds were excluded due to small numbers of records. The upper age limit was chosen to match the population who can be referred to specialist CAMHS in Scotland which is normally restricted to people aged under 18 years.

### Data

Mental health prescriptions were identified by British National Formulary (BNF) item codes recorded in PIS or by item name where there was no item code recorded. Five classes of medications have been identified as relating to mental health. The classes are defined as medications appearing in the following BNF sections: section 4.1 Hypnotics and Anxiolytics, section 4.2 Drugs used in psychoses and related disorders, section 4.3 Antidepressant drugs, section 4.4 Central Nervous System Stimulants and drugs used for ADHD and 4.10 Drugs used in substance dependence (34). These subsections align with classification of mental health medications used previously by Public Health Scotland (16) with two differences: Medications used to treat dementia were excluded as this analysis relates to children. We also included drugs normally used in the treatment of substance dependence, which are of relevance to mental health. A full list of named medications by class (BNF section) included can be found in the supplementary materials (Table S3).

Service structures to address child and adolescent mental health vary across the United Kingdom but are broadly similar. NHS Scotland provides Child and Adolescent Mental Health Services (CAMHS) for people aged under 18 and consists of multi-disciplinary teams who assess and provide treatment or interventions for young people with emotional, developmental, environmental or social factors which impact on their mental health (8). CAMHS in Scotland are structured into 4 tiers based on the type of services offered – universal and targeted services in the community (Tiers 1 & 2), locality teams based in hospital settings largely providing treatments in specialist outpatient clinics (Tier 3) and more specific services requiring additional expertise such as inpatient psychiatry (Tier 4).

All referrals to CAMHS (at tier 3) are assessed by a professional in the CAMHS team to determine if they meet the health board criteria for nature and severity of the complaint. Here, rejected referrals were defined as referrals for which the tier 3 CAMHS team declined to provide the patient further assessment or treatment in specialist outpatient CAMHS. Those who meet this definition of a rejected referral may be redirected to alternative services or provided with advice but these outcomes are not captured in the source data. Referrals were defined as accepted if the patient was assigned further assessment or treatment by CAMHS, regardless of later attendance or outcome. For analysis of accepted/rejected referrals, we excluded referrals where the outcome was still pending (i.e. they have no appointment or rejection date). This approach avoids referrals without an outcome being counted as accepted or rejected which would either under- or overestimate the proportion which are counted as rejected. Home area deprivation was measured by SIMD2020 deciles based on the recorded area of residence at the time of prescription or referral. Decile 1 is assigned to small areas ranked in the top 10% of most deprived areas in Scotland and decile 10 is assigned to the 10% least deprived areas nationally. Rates of prescription and referral have been calculated from counts (i.e. total number of prescriptions or referrals) and mid-year population estimates for people aged between 2 and 17 in NHS Grampian which are available by year, age group, sex and home-area deprivation decile (29). These population estimates are available for each year between 2015-2020 and rates for 2021 are calculated based on the 2020 population sizes.

### Data Processing

Prescribing data is missing for two months in the study period (April 2019 & December 2020) and data for 2021 is only complete until the end of May. Where average monthly prescribing rates (both prescriptions per month and monthly rate per population) for the affected years have been reported, these are calculated from the total count available divided by the number of months in the year with complete data. Where monthly counts have been calculated into rates and reported (e.g. by BNF section in Figure 2) the two missing monthly totals have been imputed based on the previous monthly total.

From mid-2019 to mid-2020 there is a marked drop in the number of prescriptions in total, and specifically for the ‘Hypnotics and Anxiolytics’ BNF section (see Figures 1 & 2). Most prescriptions in this section both before and after this time period are for melatonin which has some licensed and some unlicensed preparations. Unlicensed preparations of prescribed medications are not indexed in the PIS pricing codes and are thus not recorded in the PIS system which means it is not possible to determine if this drop reflects a true drop in melatonin prescribing.

**Figure 1.**
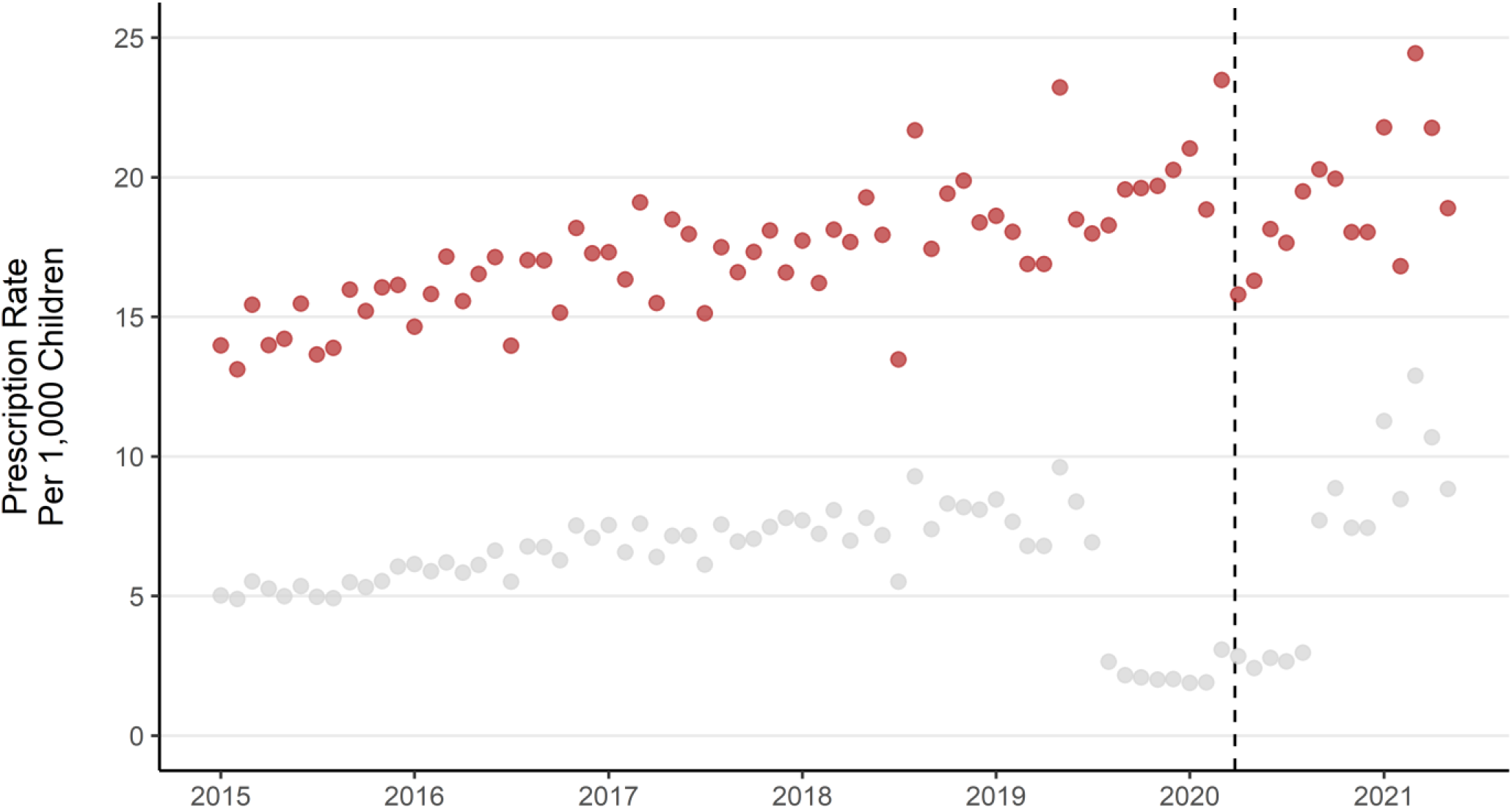
Monthly mental health prescription rate per 1,000 children aged 2-17. Dashed line indicates the first UK COVID-19 lockdown beginning 2020/03/23. Grey points for melatonin prescriptions only, Red for all prescriptions excluding melatonin.

**Figure 2.**
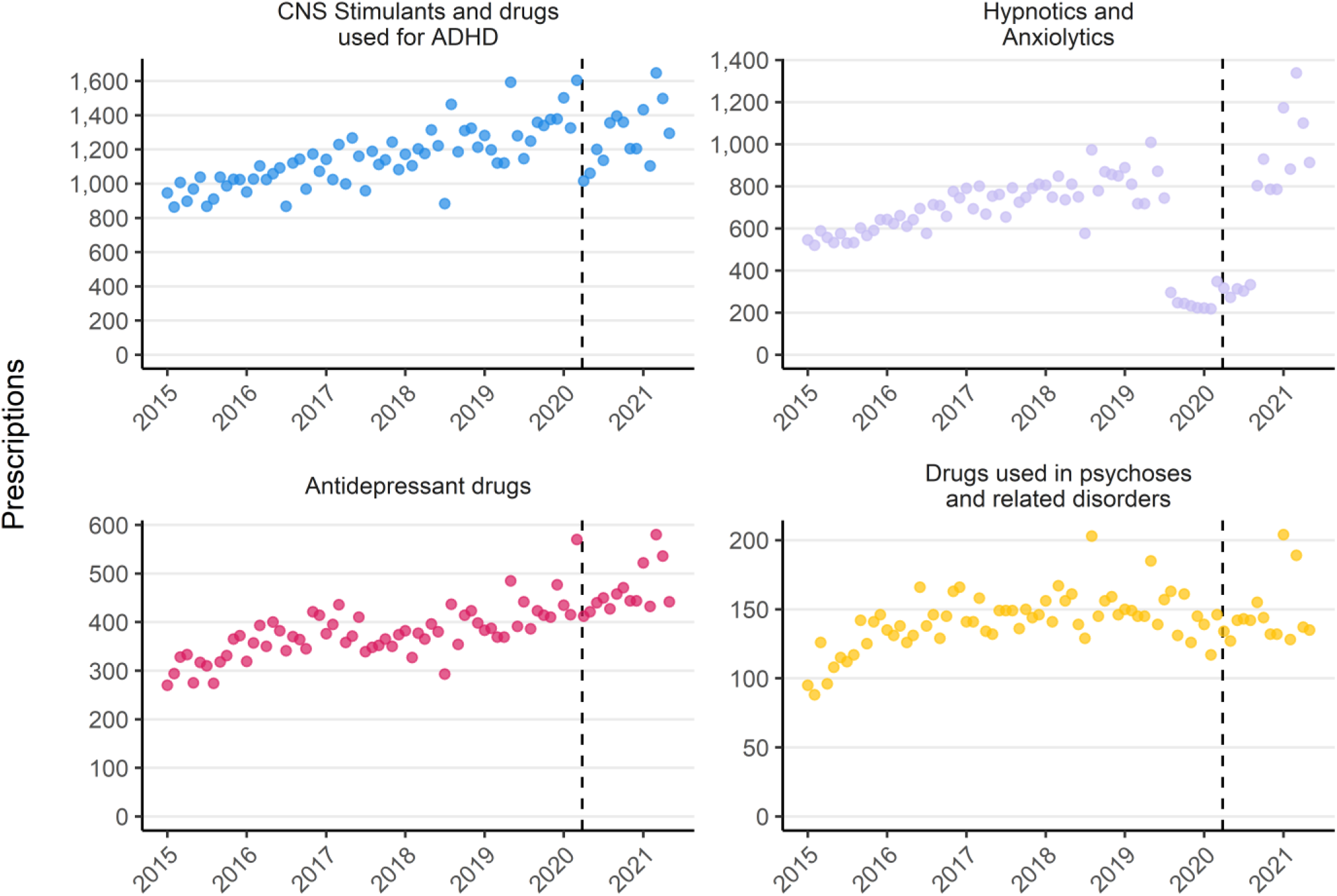
Monthly mental health prescriptions by BNF Section (January 2015 – May 2021). Dashed line indicates the first UK COVID-19 lockdown beginning 2020/03/23. Note: Y axis scale varies and drugs for treatment of substance dependence are not shown due to low monthly counts

### Statistical Methods

The primary outcomes examined were counts, rates and proportions of the population who received mental health prescriptions or specialist CAMHS referrals (both accepted and rejected). Secondary outcomes evaluated were differences between these counts and rates by age, sex and deprivation

Relative and absolute inequality gaps in rates of mental health prescription and specialist CAMHS referral by area deprivation were quantified by calculating Slope Index of Inequality (SII) and Relative Index of Inequality (RII) using the linear regression approach (35). SII is an absolute measure which estimates the difference between rates in the most and least deprived groups, accounting for differential population size. RII is a relative measure which estimates the difference between rates in the most deprived group compared with the mean rate in the population (36). Both SII and RII have been calculated for annual rates and for the mean of annual rates over the whole study period.

## Results

### Overview

The average annual population size between 2015 and 2020 for children aged 2-17 in NHS Grampian was 99,591. The average annual proportion of children with at least one mental health prescription was 3.1% and with a specialist CAMHS referral was 2.8%. There was a total of 178,657 mental health prescriptions for 8,170 children (Table 1) and 21,874 CAMHS referrals for 15,022 children (Table 2) over the study period. Period prevalence of mental health prescription was 8.2% (2015 to May 2021) and for specialist CAMHS referral was 15% (2015 – October 2021). A total of 18,732 unique individuals were identified across prescribing and referrals datasets, with 4,460 (24%) appearing in both. 3,710 children had a prescription but no referral (45% of all with a prescription) and 10,562 had a referral but no mental health prescription (48% of all with a referral). The mean annual number of prescriptions was 25,522 (for an average of 3,093 individuals) and the mean annual number of specialist CAMHS referrals was 3,125 (for an average of 2,823 individuals).

**Table 1.**
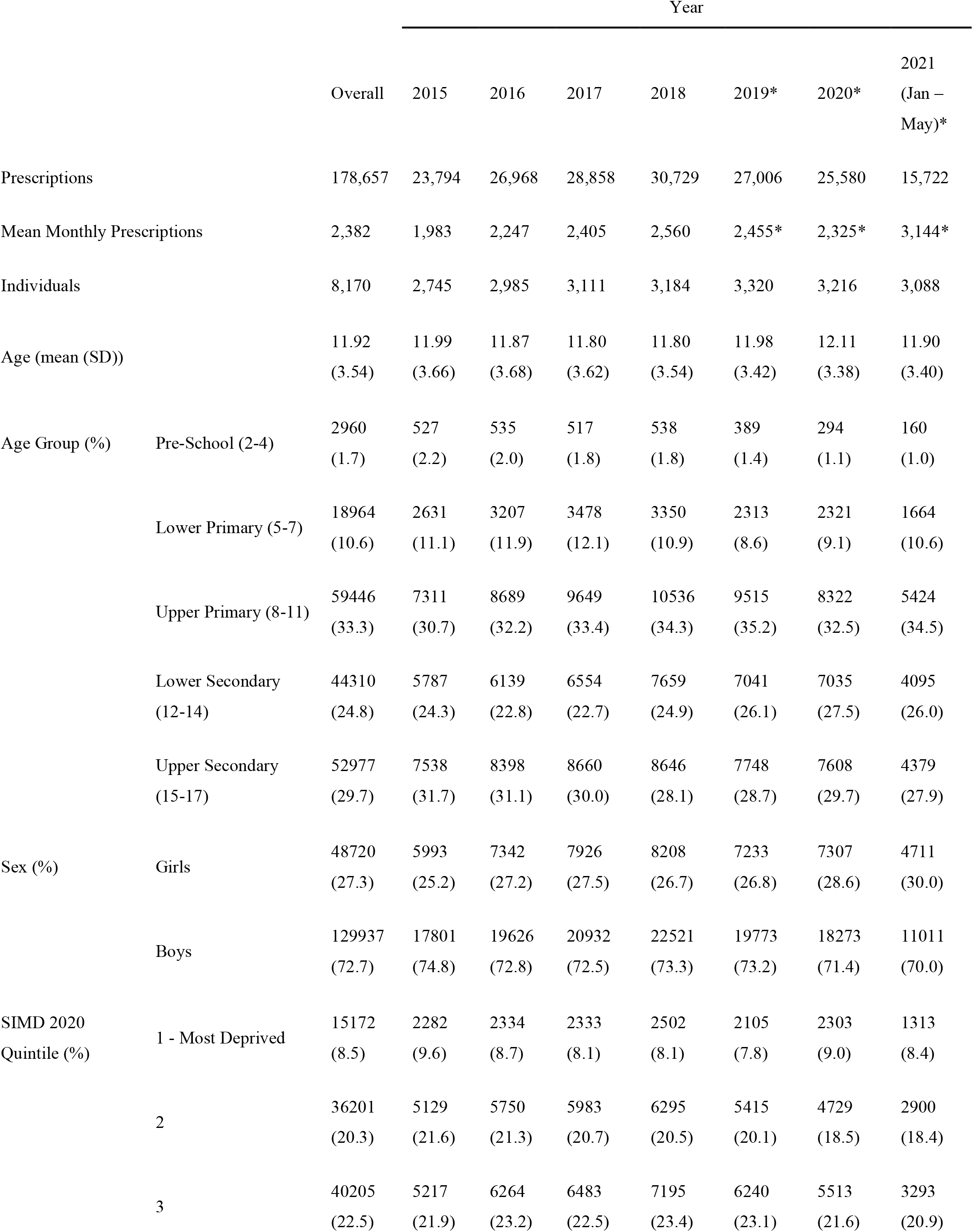
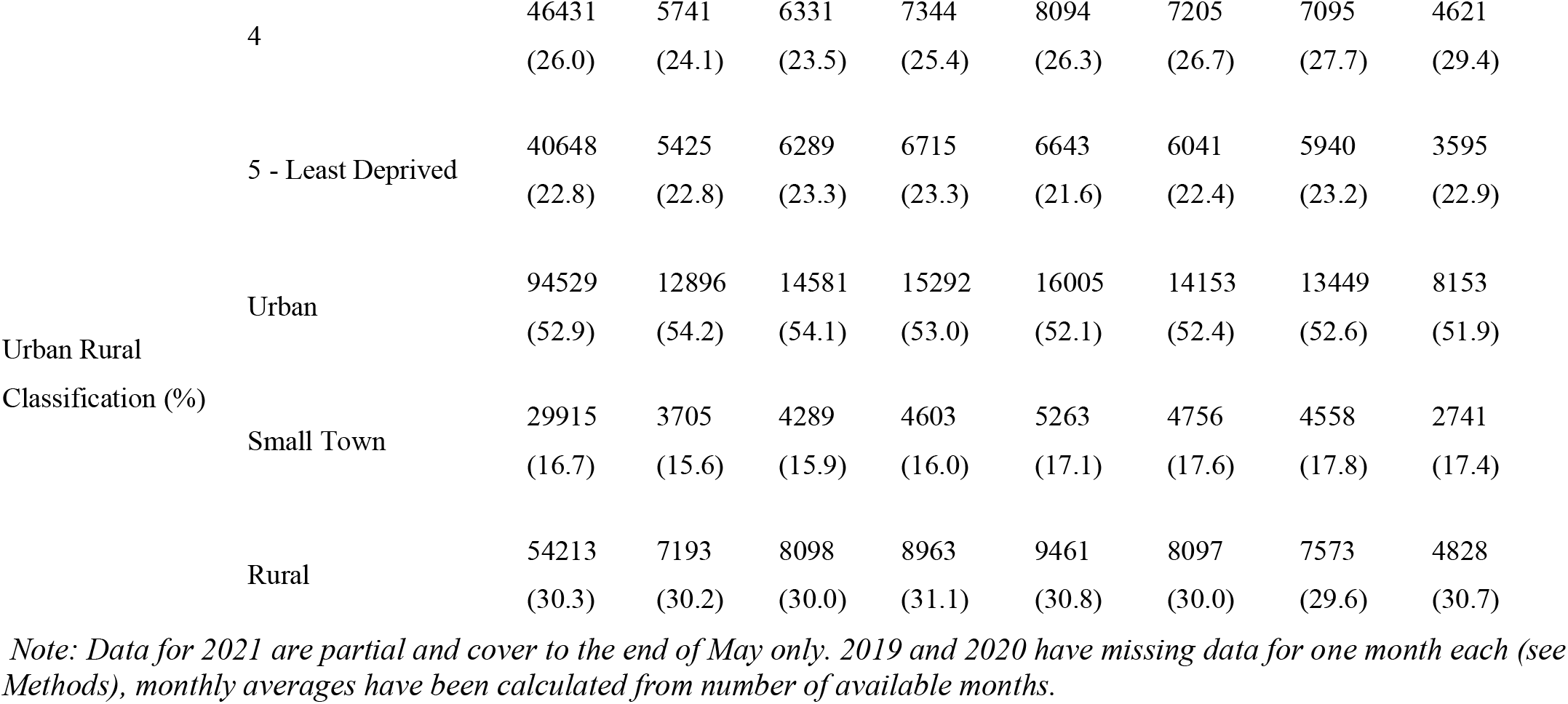
Summary of mental health prescriptions for children.

**Table 2.**
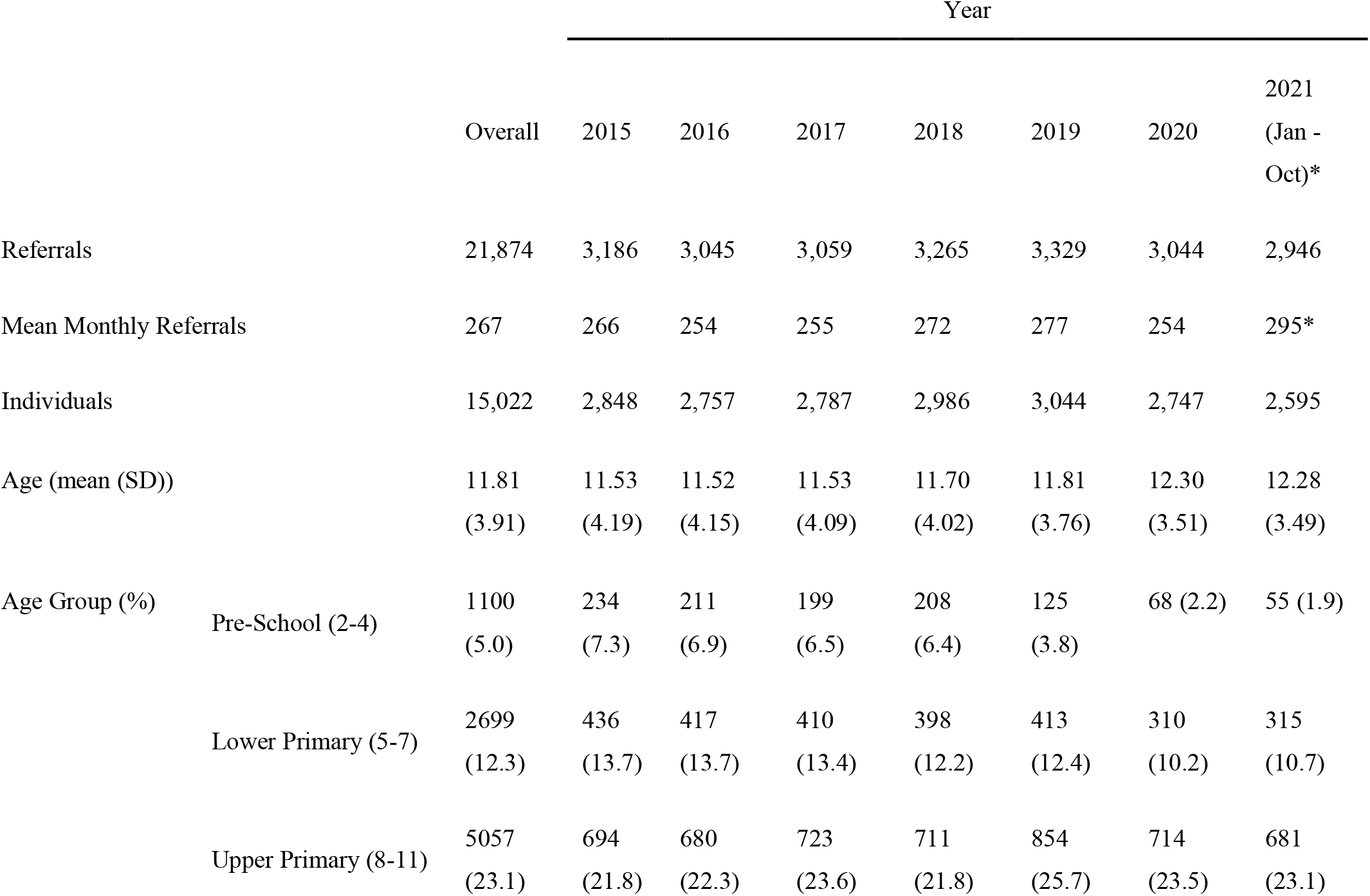
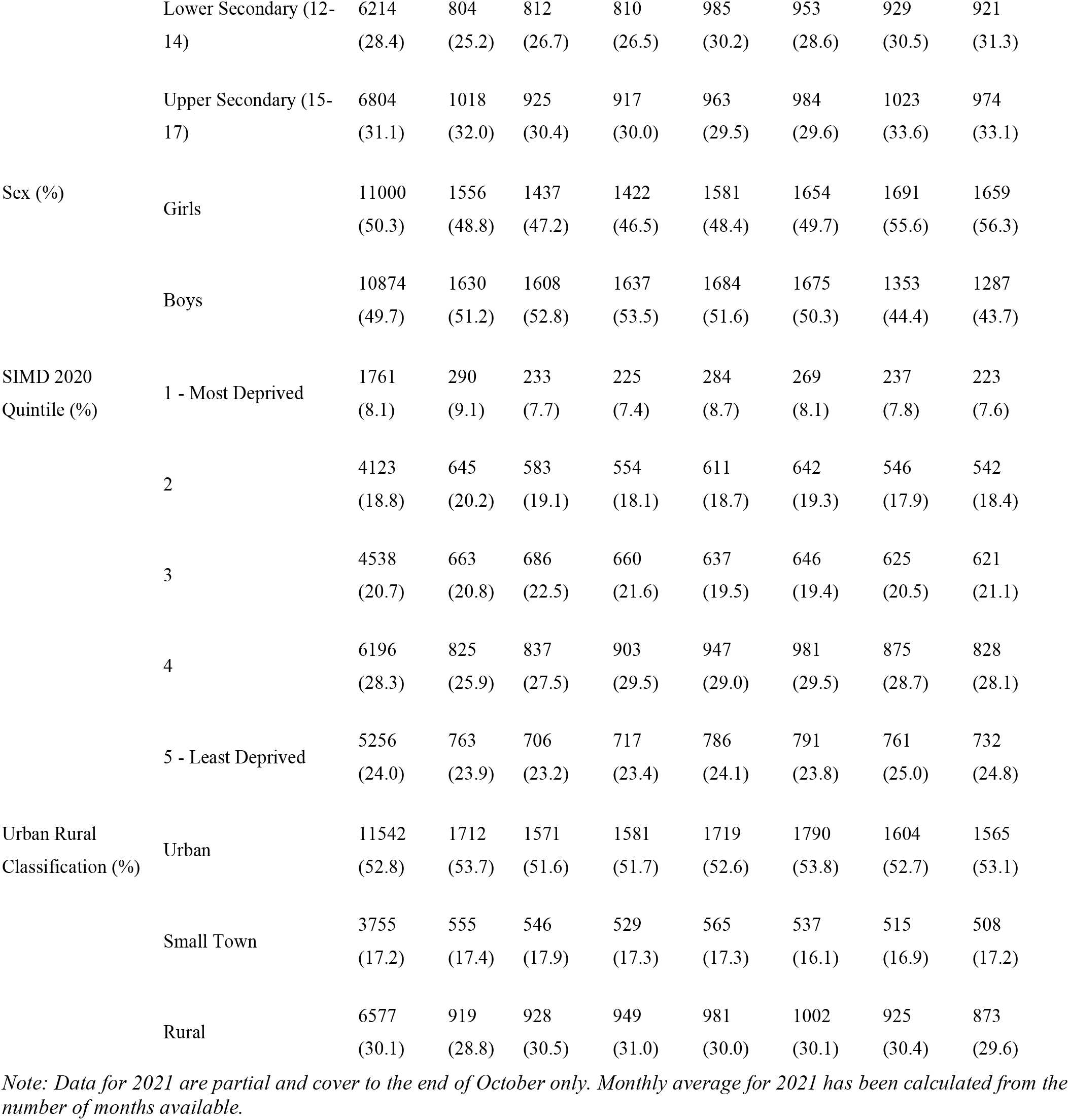
Summary of referrals to specialist CAMHS.

### Mental Health Prescriptions

#### General Trends

The monthly rate of mental health prescriptions (for any medication listed in supplementary table S3) per 1,000 children has risen by 56% over the study period, from an average of 20 per month in 2015 to 31 per month up until the end of May in 2021. A drop in monthly rate of prescription can be seen at the start of the 1^st^ UK COVID-19 lockdown in March 2020. There is a prolonged drop in the rate of prescriptions for melatonin specifically from August 2019 to August 2020 (Figure 1) which represents either a real drop in the number of prescriptions during this period, or that prescriptions for melatonin were filled with preparations which are not recorded as billable items in the PIS dataset (S. McTaggart, personal communication, 14^th^ September 2021). Supplementary figures S1-S5 are replications of figures 2-6 in the main body of this manuscript using data which excludes all melatonin prescriptions.

Rises in the rate of prescription are observed for each class of medication apart from treatments of substance dependence (Figure 2). Prescriptions of medications to treat anxiety and sleep disorders (Hypnotics and Anxiolytics) have increased 91% from 565 per month in 2015 to 1,082 in 2021. When melatonin is excluded, prescriptions in this class reduced between 2015-2021 (Figure S1). Antidepressant prescriptions have risen by 59% (316 monthly in 2015, 502 in 2021), ADHD medications have risen by 45% (965 in 2015, 1,395 in 2021) and drugs used for psychoses and related disorders have risen by 35% (118 in 2015, 159 in 2021), although most of this rise occurred between 2015 and 2016 and the subsequent trend is not clear.

The monthly rate of mental health prescription per 1,000 children has increased over the study period in each age group apart from the youngest (Figure 3). Rates of prescription have consistently been higher in older age groups. The largest increase over the period was for children in the lower primary school age group, which rose by 60% from an average of 10 prescriptions per month per 1,000 children in 2015, to 17 per month in 2021. The rate of prescription in pre-school aged children dropped by 22% from an average of 2 per month per 1,000 children in 2015 to 1.7 per month in 2021. Excluding prescriptions for melatonin did not dramatically change trends by age (Figure S2).

**Figure 3.**
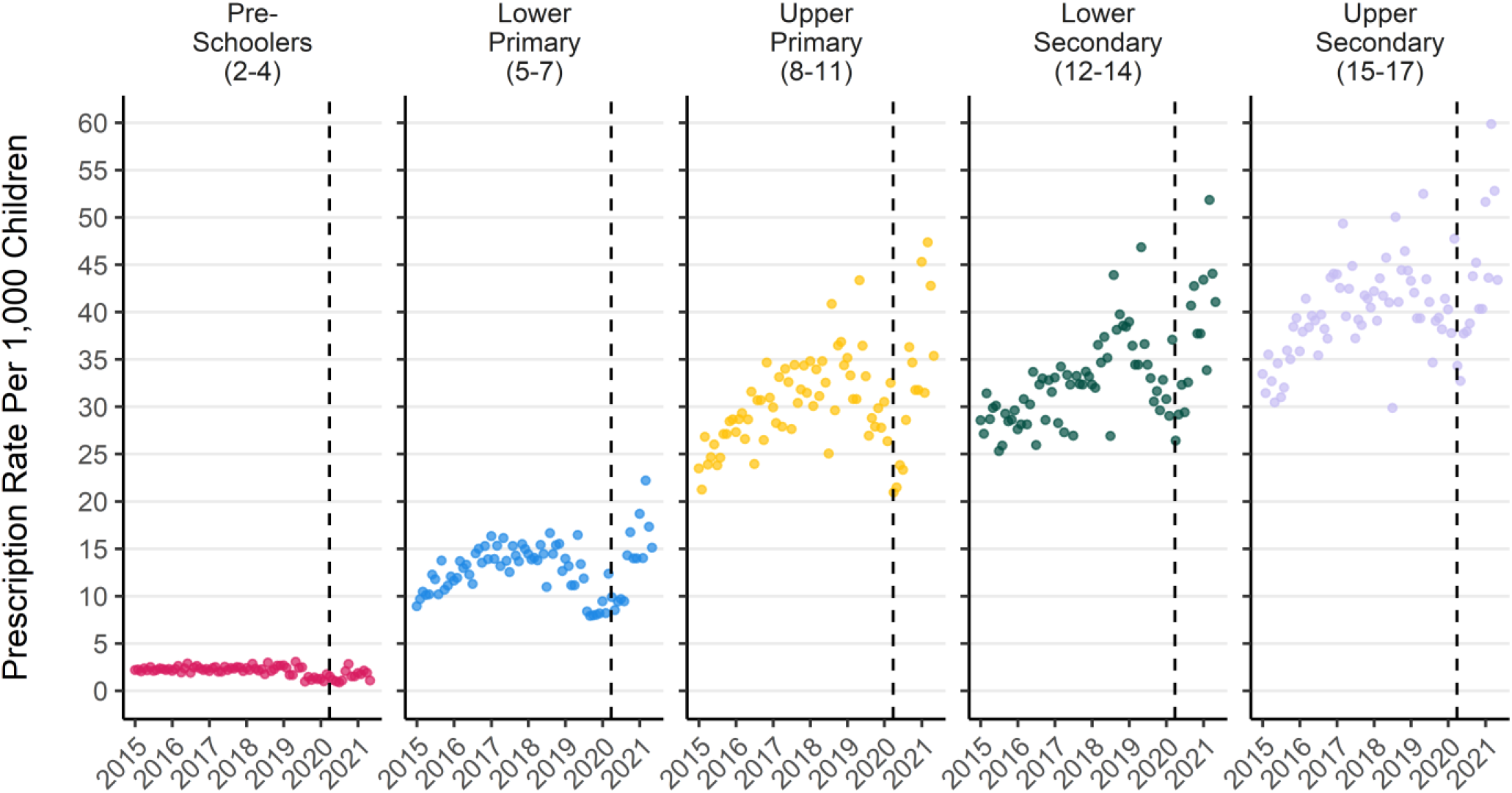
Monthly mental health prescription rate per 1,000 children by school age group. Note: the drop in melatonin prescriptions recorded from mid 2019 to mid 2020 is more pronounced in younger ages. Dashed line represents the start of the 1st UK COVID-19 lockdown.

The monthly prescription rate per 1,000 children increased for both boys and girls over the study period (Figure 4a). Boys had consistently higher rates of prescription over the study period, but girls saw the larger increase. The monthly rate of prescription per 1,000 children for boys rose by 45% from an average of 29 per month in 2015 to 43 in 2021. For girls the rate was 10 per month in 2015 and rose by 84% to 19 per month in 2021. The drop in melatonin prescriptions had a more marked effect on monthly prescription rates for boys than girls in 2019 and 2020 but does not affect the 2021 average rate. Overall trends of increasing prescriptions for both boys and girls, but larger increases for boys were also evident when melatonin prescriptions were excluded (Figure S3a)

**Figure 4.**
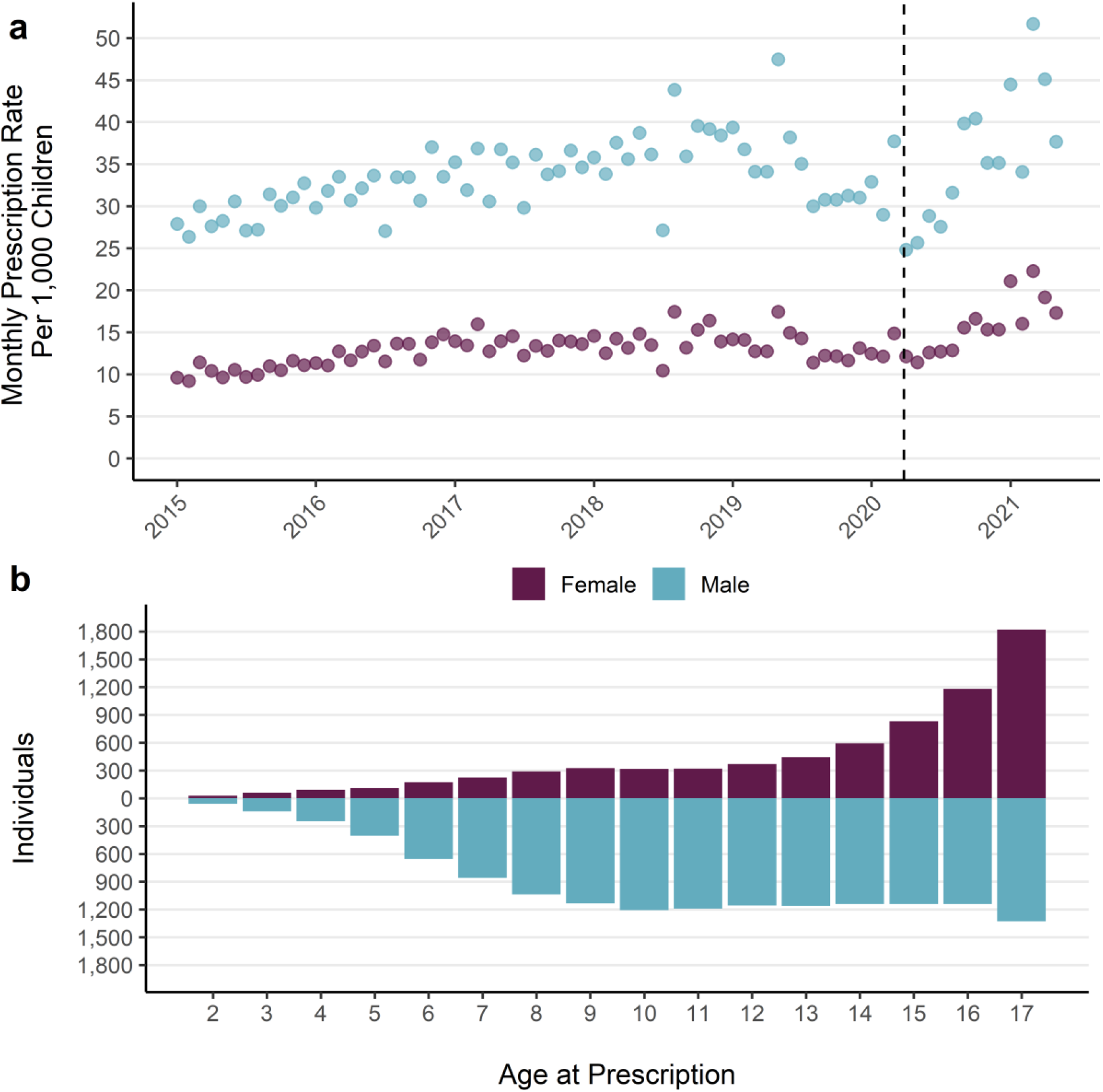
a) Monthly prescription rate per 1,000 children by sex. b) Count of unique individuals with a mental health prescription by age at prescription. Note: includes period of lower melatonin prescribing which is more prominent for boys.

### Group Differences

Over the whole study period boys accounted for 73% of total prescriptions and 57% of unique individuals with a prescription. The overall rate of prescription for boys (28 per person) was double that of girls (14 per person). Prescriptions for girls are mostly concentrated in older ages and the distribution of prescriptions generally increases with age (Figure 4b). The distribution of age at prescription for boys is more evenly spread than for girls, with a larger number of individuals receiving a prescription at all ages apart from the oldest ages (16 and 17 years) where more girls received prescriptions. Gender differences in prescribing are consistent over the study period and mental health prescribing has increased for both boys and girls (Figure 4a) although the increase was larger for boys. These patterns are maintained when prescriptions for melatonin are excluded (Figure S3a and b).

The medicines with the most prescriptions over the whole study were used to treat ADHD (top drug: methylphenidate hydrochloride) and were more common for boys and in younger age groups. Medications to treat ADHD accounted for more than half of all mental health prescriptions for children in upper primary and lower secondary school (Figure 5). The medicines prescribed to the most individuals was medication for sleep disorders (top drug: melatonin) which are mostly prescribed in younger ages. Antidepressants had the second-most individuals who received a prescription with 3,427 over the study and they are the most prescribed for upper secondary aged children (Figure 5). Excluding prescriptions for melatonin dramatically reduces the share of medications in the hypnotics and anxiolytics class in all age groups (Figure S4), with treatments for ADHD becoming the most common and accounting for 90% of prescriptions for those aged between 2-11 years.

**Figure 5.**
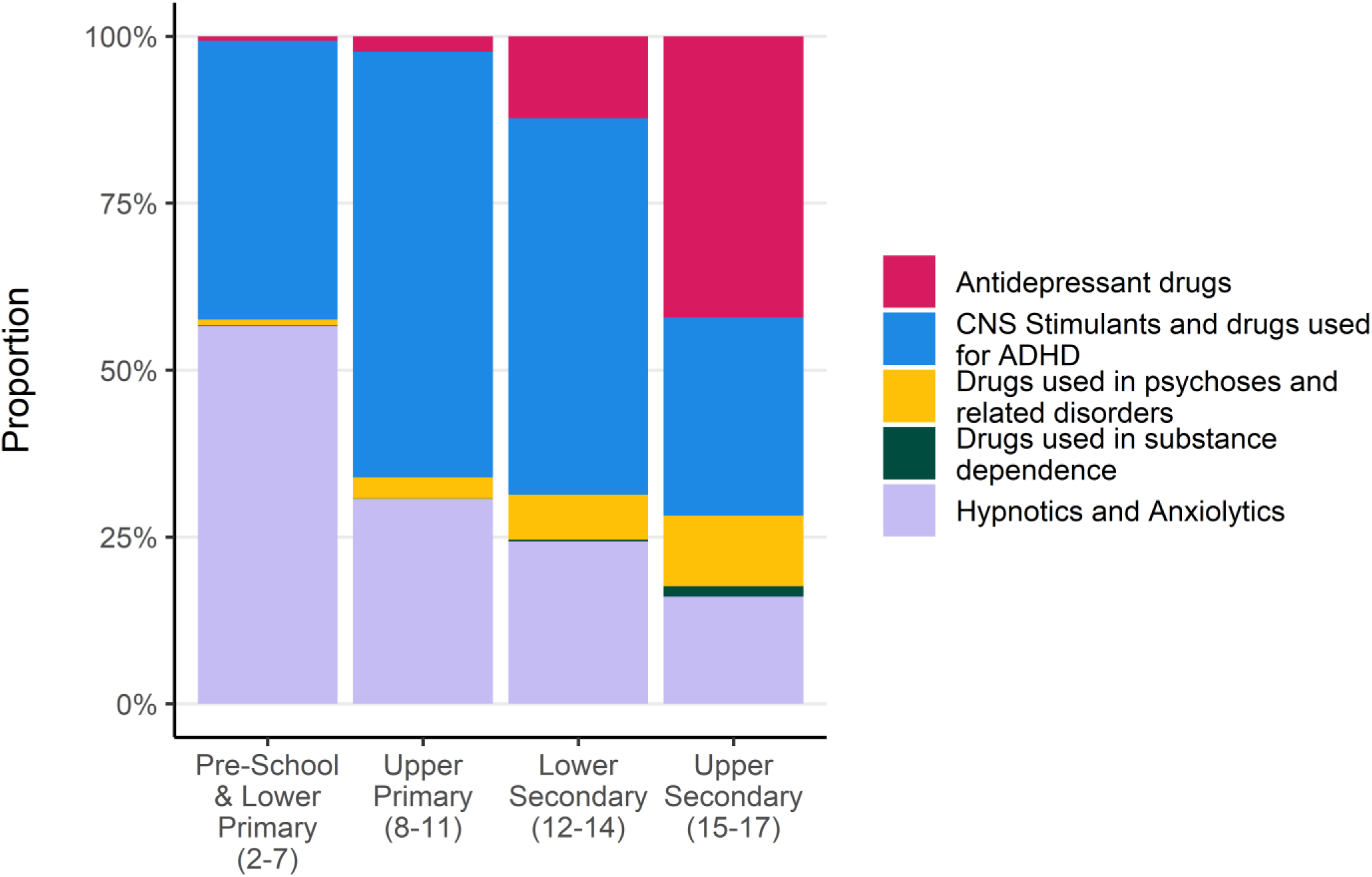
Proportion of total prescriptions by BNF Section and school age group

### Inequalities by Area Deprivation

There is a clear social gradient in mental health prescribing by home-area socioeconomic deprivation. Over the whole study period, the average annual rate of prescription for children living in the most deprived 10% of neighbourhoods was double that of the least deprived, with 34 prescriptions per 100 children in the most deprived areas and 17 per 100 in the least deprived. The average annual proportion of the population in the most deprived 10% of neighbourhoods receiving a mental health prescription was 4%, which was double the proportion in the least deprived decile at 2%. The mean age at first prescription for those in the most deprived decile is 11.9 years (SD: 4.4), almost 11 months younger than those in the least deprived decile (Mean: 12.8 years SD: 3.8).

The social gradient was consistent over the study period, with those in the most deprived decile receiving around 23 additional prescriptions per year per 100 children compared with those in the least deprived decile (Figure 6a). Relative to the average for the whole population the annual rate of mental health prescription in the most deprived area was between 40-50% higher (Figure 6b). Inequalities in prescription rates by area deprivation remained when prescriptions for melatonin were excluded (Figure S5a and b).

**Figure 6.**
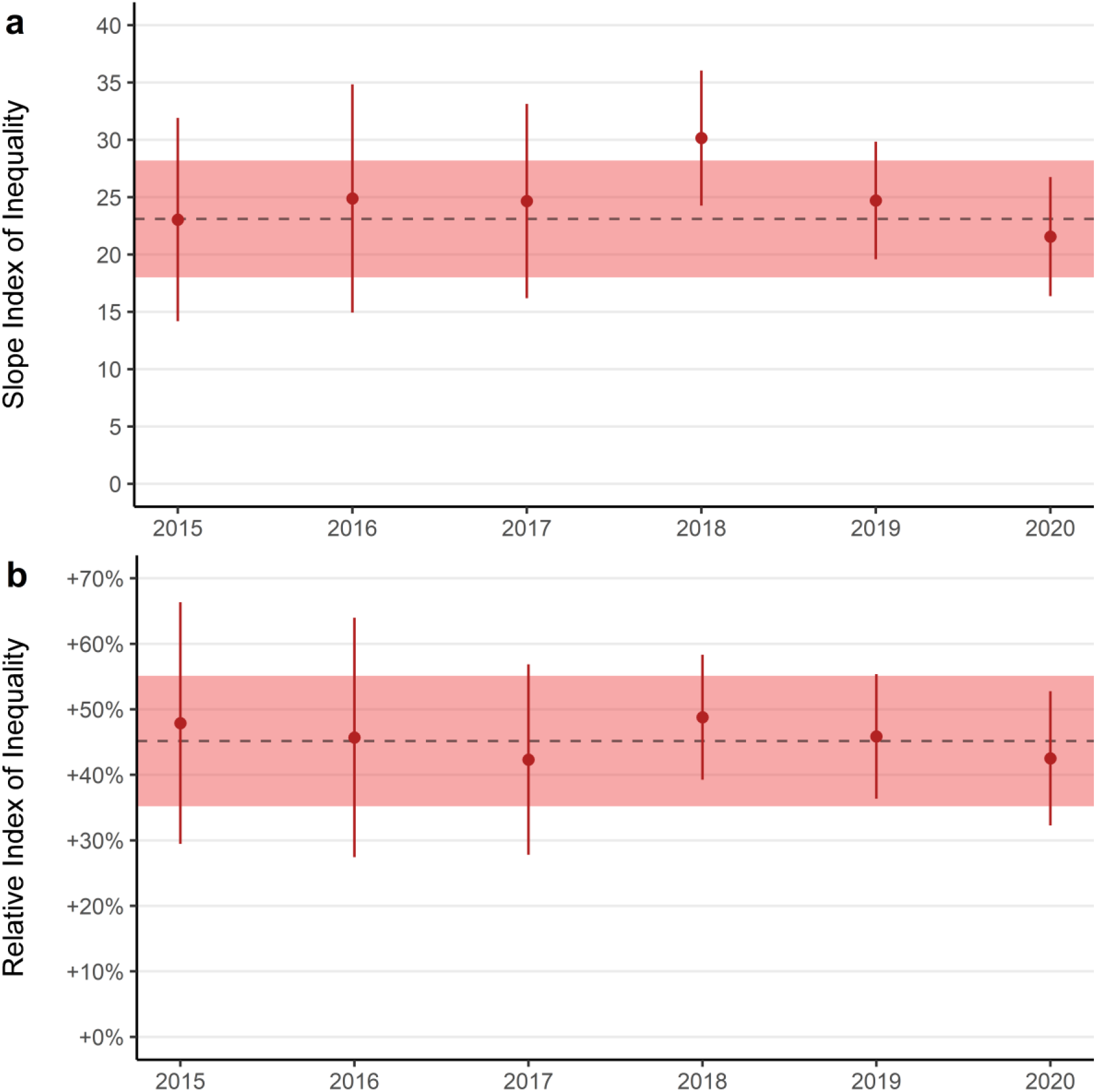
a) Slope Index of Inequality and b) Relative Index of Inequality for annual mental health prescriptions per 100 children by area deprivation. Point ranges denote annual SII and RII with confidence intervals. Dashed lines denote mean annual SII and RII over the entire study period and the shaded red area represents the mean annual confidence intervals.

### Specialist CAMHS Referrals

#### General Trends

The rate of referrals to specialist CAMHS rose by 9% from 32 per 1,000 children per year in 2015, to 35 in 2021. In 2015 the mean number of referrals to specialist CAMHS per month was 266, which rose by 11% to a mean of 295 through October in 2021. The increase in monthly referrals has mainly occurred in the period following the first UK COVID-19 lockdown in March 2020, after an initial drop in referrals (Figure 7a).

**Figure 7.**
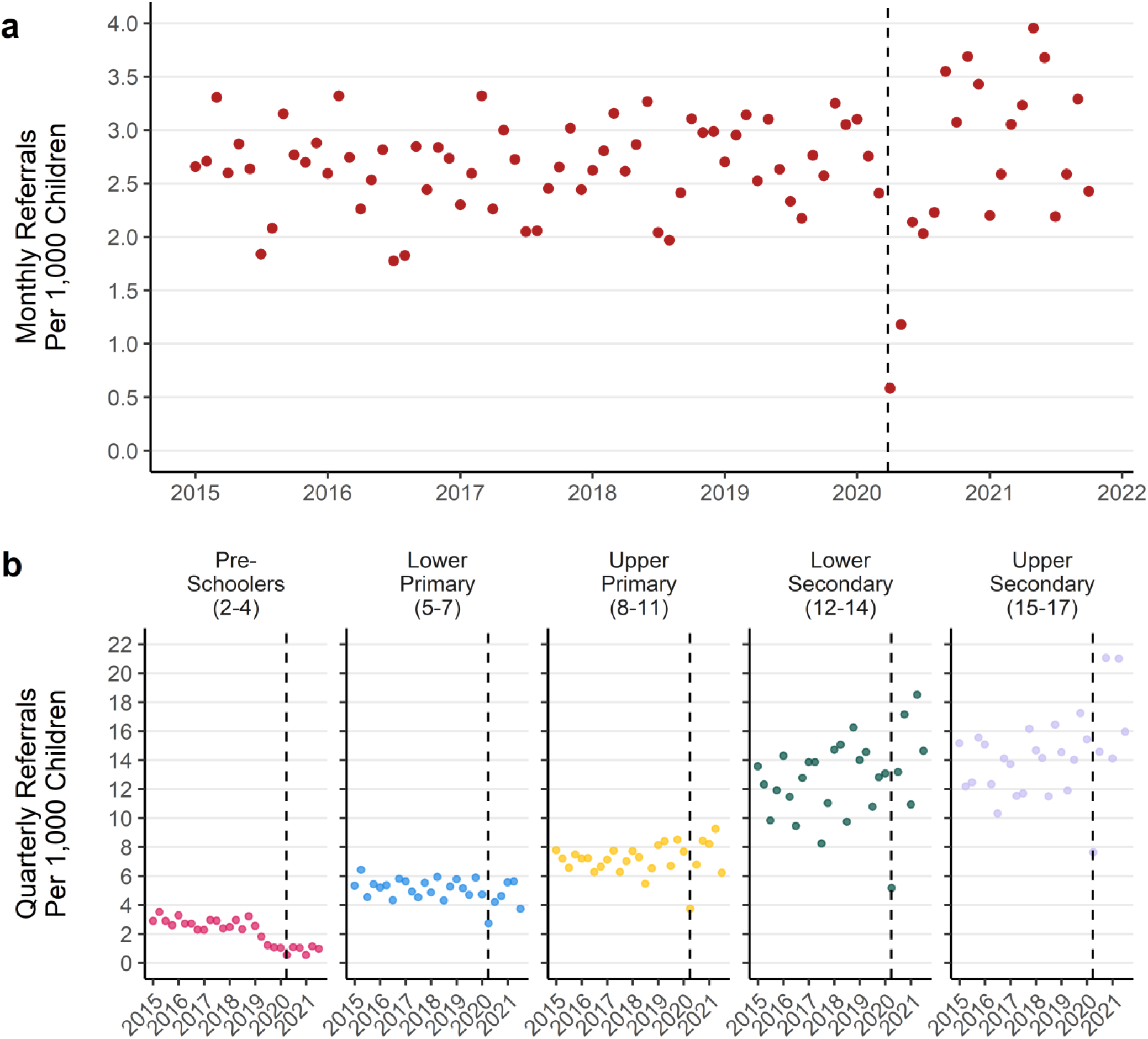
a) Monthly rate of referrals per 1,000 children b) Quarterly referral rate per 1,000 children by age group (January 2015 to October 2021)

There were different patterns of referral through time by age group, with decreasing referrals for younger children and increasing referrals for older age groups (Figure 7b). Children in the youngest age group had a 70% reduction in the rate of referrals per quarter from an average of 2.9 per 1,000 children per quarter in 2015, to 0.9 per 1,000 per quarter in 2021. For the oldest age group this number increased by 23% from an average of 14 per 1,000 per quarter in 2015 to 17 per 1,000 per quarter in 2021.

#### Group Differences and Change

The monthly specialist CAMHS referral rate was the same between boys and girls until the start of the first UK COVID-19 lockdown, after which there was an increase for girls but not for boys (Figure 8a). Both boys and girls had average monthly referral rate of 2.7 per 1,000 children in 2015 which rose by 25% for girls in 2021 to 3.4 per 1,000 per month and fell 6% for boys to 2.5.

**Figure 8.**
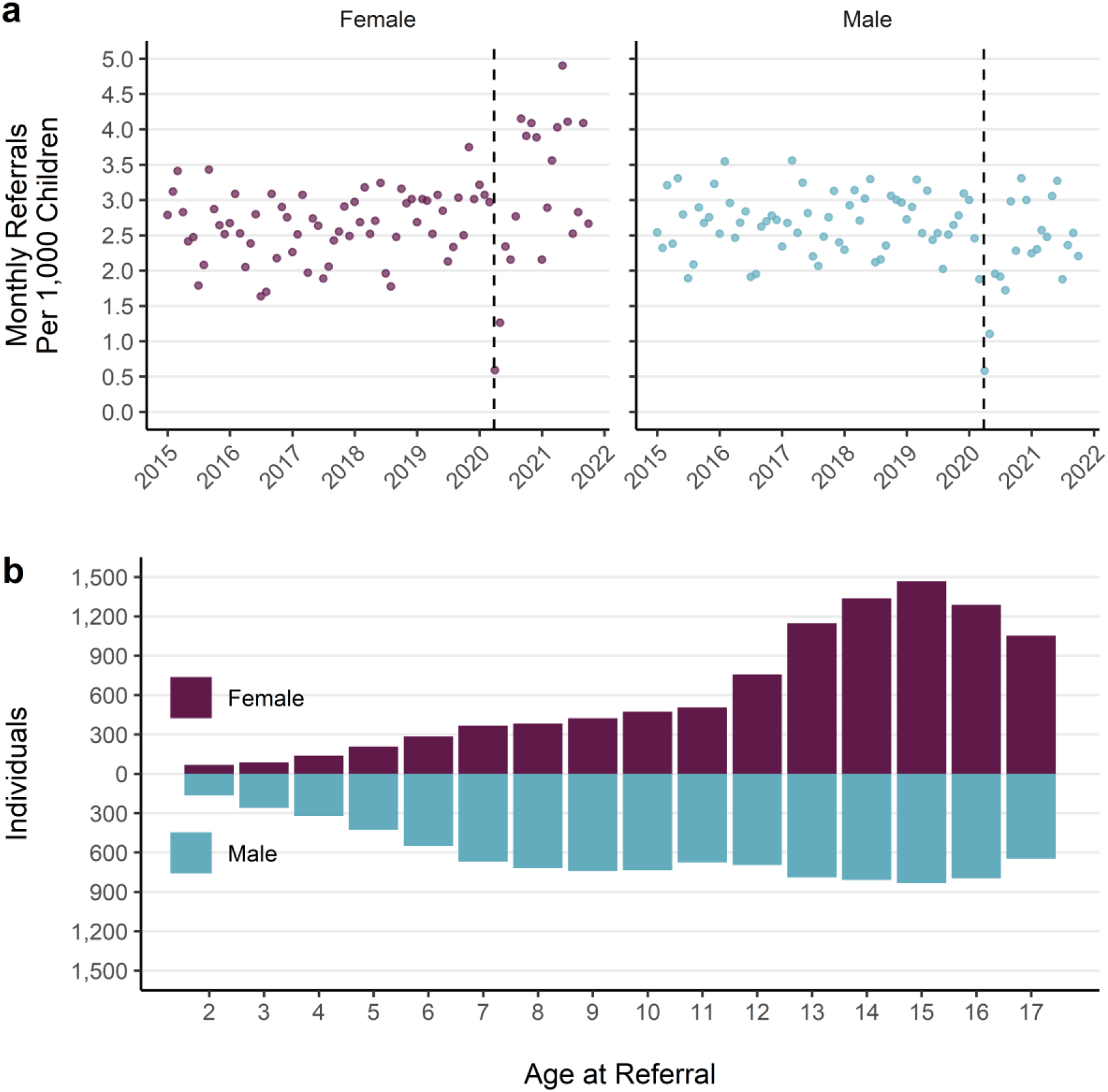
a) Monthly referral rate per 1,000 children by sex. b) Distribution of age at referral for unique individuals by sex

The distributions of age at referral differ for boys and girls (Figure 8b). For boys, referral rates are higher at younger ages and remain steady through the teen years. Boys have more individuals with a referral at each year of age than girls up until 11 years. Referrals for girls follows a different pattern, with growing numbers of individuals referred as age increases, until 12 when there is a rapid rise in referrals.

#### Inequalities by Area Deprivation

Over the entire study period, there is a gradient in referral rates by home-area deprivation level. The rate of referral for children living in the most deprived areas is 1.9 times that in the least deprived areas. The social gradient has been consistently high over the study period. The most deprived areas see between 2 and 2.5 additional referrals per 100 children each year (Figure 9a). The rate of referral in the most deprived areas has been between 30-40% higher than the average of the whole population each year (Figure 9b). The mean age at which children in the most deprived areas are first referred to specialist CAMHS is 10 years and 5 months, over a year younger than the average in the least deprived areas (mean: 11 years and 8 months).

**Figure 9.**
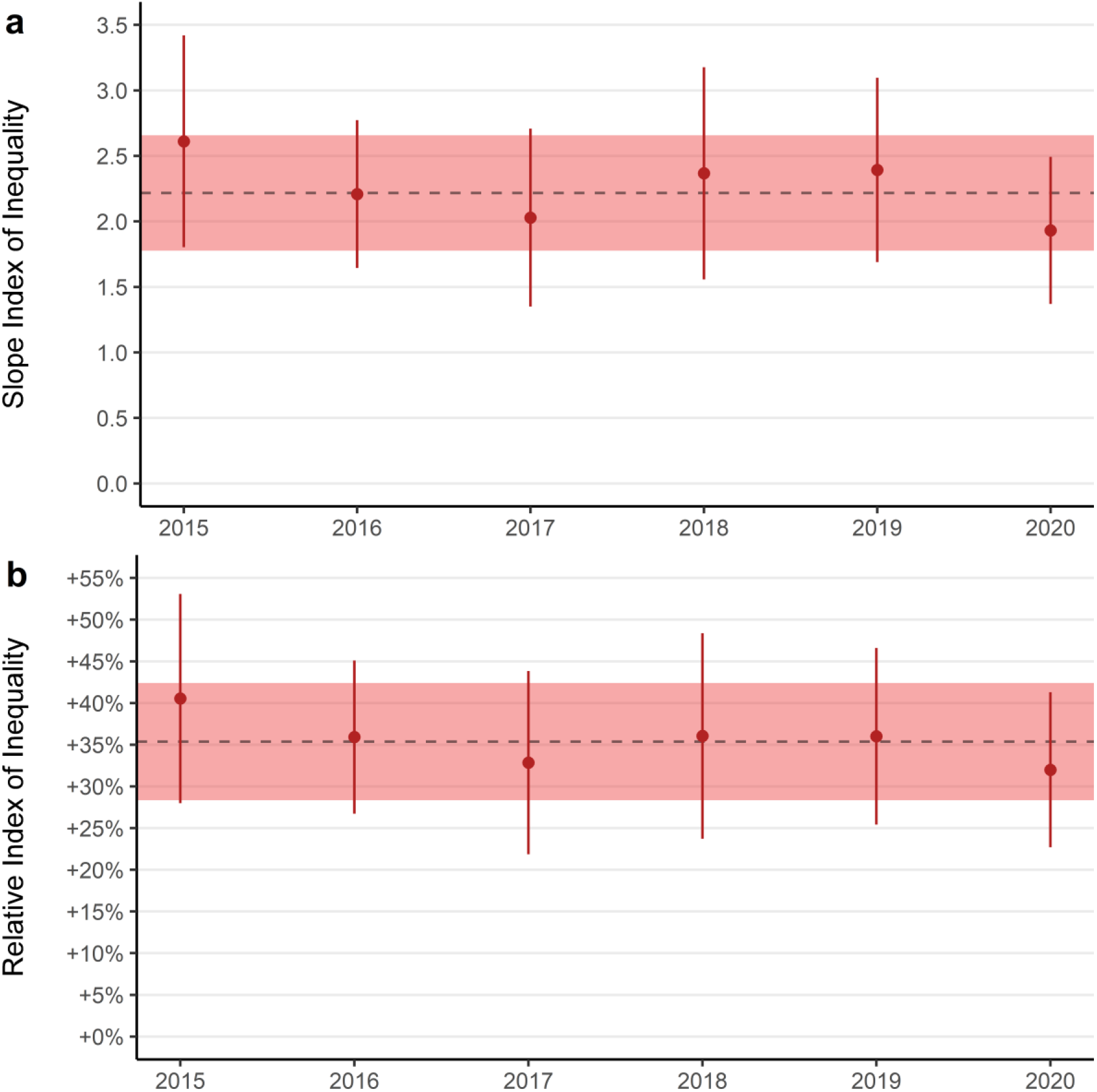
a) Slope Index of Inequality and b) Relative Index of Inequality for annual specialist CAMHS referrals per 100 children by area deprivation. Point ranges denote annual SII and RII with confidence intervals. Dashed lines denote mean annual SII and RII.

#### Rejected Referrals

The proportion of referrals to specialist CAMHS which were rejected nearly doubled from 17% in 2015 to 30% in 2021 (Figure 10a). The proportion of referrals which were rejected increased following the first UK COVID-19 lockdown (Figure 10a). Characteristics of people with rejected referrals also changed through time. The proportion of referrals for boys that were rejected increased throughout the study period. In contrast, the proportion of referrals for girls that were rejected remained roughly static until after the 1^st^ COVID-19 lockdown when their rejection rate rose (Figure 10b). Between 2015 and the end of 2017 rejection rates were between 10% and 20% for all age groups. Since 2018, the proportion of referrals being rejected in pre-school aged children rose to 67% in 2021 while rejection rates for secondary school aged children remained steady at roughly 20% (Figure 10c).

**Figure 10.**
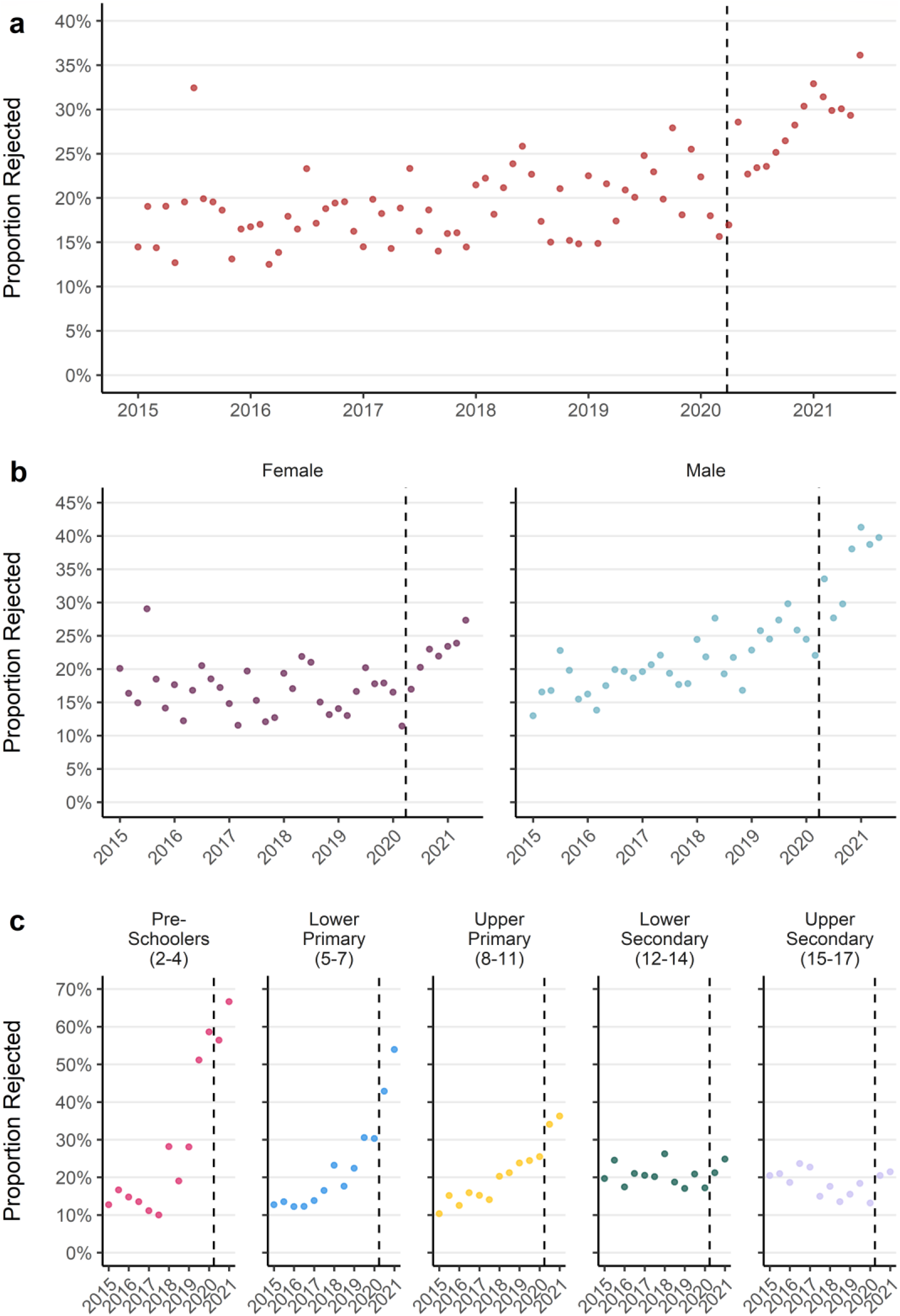
a) Monthly proportion of referrals rejected b) Monthly proportion of referrals rejected by sex c) Quarterly proportion of referrals rejected by age group

### Accepted Referrals

Changes in the populations receiving referrals and rejections has changed the characteristics of who was able to access specialist CAMHS treatment over the study period. The proportion of new patients receiving specialist outpatient treatment who were girls increased from an even standing with boys in the early part of the study period, to almost two-thirds girls each month in 2021 (Figure 11). Since January 2019 there has only been one month in which more accepted referrals were for boys. In addition, the mean age of children whose referrals were accepted increased over the study period, from 11 years and 4 months (SD: 4.2) in 2015 to 12 years and 10 months (SD: 3.2) in 2021.

**Figure 11.**
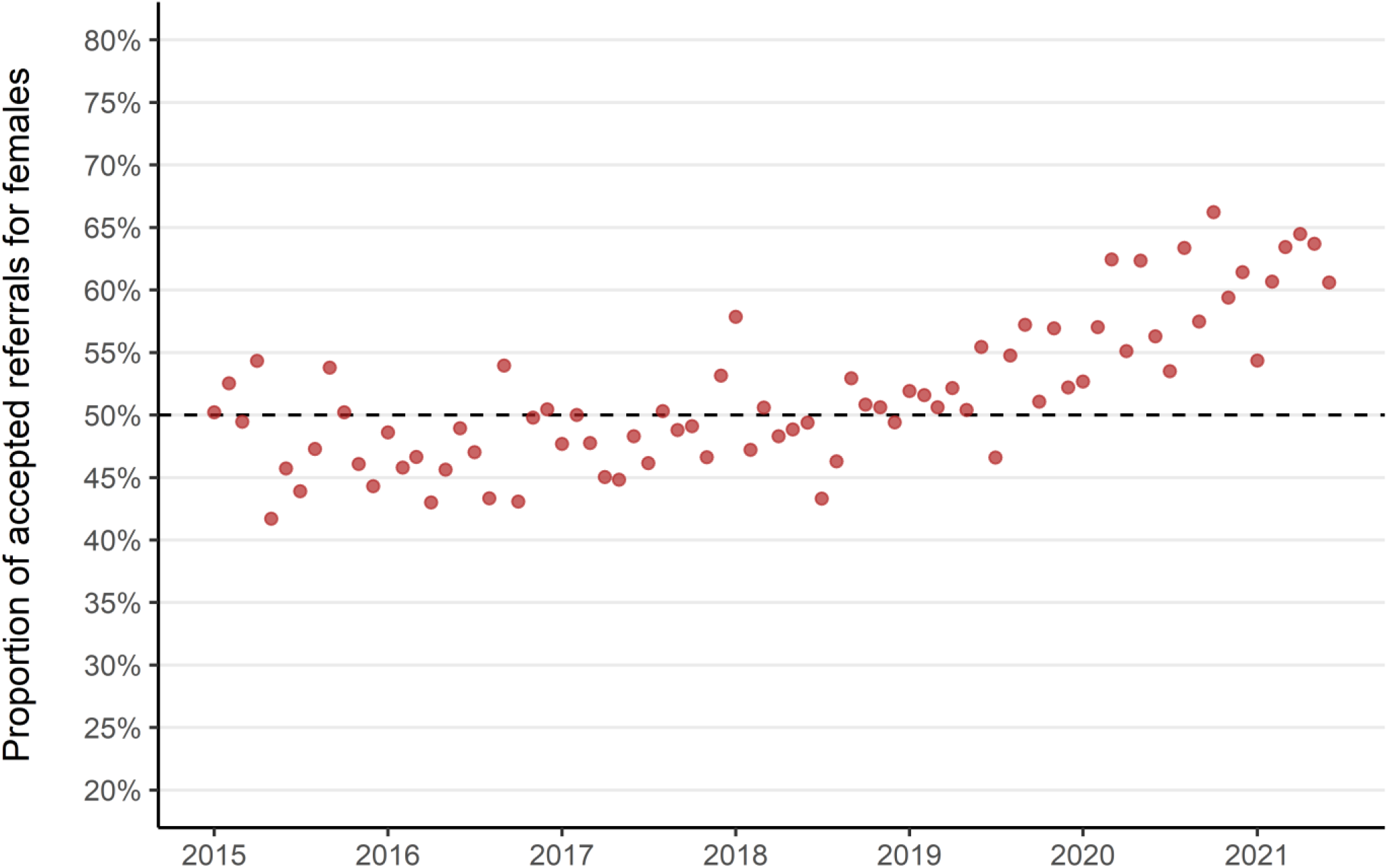
Monthly proportion of accepted referrals which were for girls (2015 - 2021). Dashed line indicates even referral acceptance for boys and girls.

## Discussion

### Principle Findings

We found a substantial increase of mental health prescribing in children across medicines used to treat a variety of mental health conditions – with rates of anti-anxiety and sleep medication prescriptions increasing by 90% and anti-depressants by 60%. There are stark differences in who is receiving prescriptions for mental health drugs – in younger children prescriptions are predominantly for boys to treat ADHD, and in older children prescriptions are predominantly for girls to treat depression.

Overall, we found that referrals to specialist CAMHS were stable until the 2020 COVID-19 lockdown, after which there was an increase particularly among girls and those in older age groups. We found that the proportion of referrals which were being rejected increased before the COVID-19 pandemic, then accelerated following the first 2020 lockdown. The increase in rejected referrals has had the biggest impact in the youngest ages and for boys. This has resulted in a static number of accepted referrals but a change in the characteristics of the treated population who are more likely to be girls and in older age groups.

We also found a clear and persistent social gradient by area deprivation for rates of both mental health prescribing and specialist CAMHS referral. Rates for both prescribing and referrals to specialist care for children living in the most deprived areas were twice as high compared with the least.

### Strengths and Limitations

The data used in this study are comprehensive for the entire population of children in the NHS Grampian region. The Prescribing Information System database is the definitive data source for medicines prescribed and dispensed in the community in Scotland. The CAMHS referrals data used in this study are required to be recorded locally by the Scottish Government for quarterly reporting of national statistics on patient waiting times. This work adds value to these high-quality administrative healthcare records through individual level linkage of socioeconomic and demographic information. Investigation of the individual level records also reveals trends which are not captured in aggregate statistics available as open data. This detailed descriptive analysis will support future research aiming to identify hypotheses for potential mechanisms which may explain these findings, inform service planning and policies to provide treatment for equitably. This work was also informed by public involvement and engagement activities at each stage, including in research design, analysis methods and interpretation of results. A full description of these activities is included in the supplementary materials (Table S1).

The data available to this study are of a high quality but do not represent the totality of mental health care services which are available to children and young people, although unmeasured/alternative sources of treatment for mental health (e.g. lower tier community-based CAMHS or private and charitable organisations) are mostly aimed at people with lower severity needs. Due to the administrative nature of these data sources there is very little information relating to clinical indications. In the case of prescribing data, although medicines are grouped in the BNF based on their common usage, some medications are indicated for the treatment of a variety of conditions or can be used ‘off license’. This means that we are unable to determine conclusively why any given prescription has been given to a patient. For CAMHS referrals, additional information was not available about the clinical complaints of those receiving referrals, or for those whose referrals are rejected. It is therefore not possible to determine the reason for a rejected referral or whether those whose referrals are rejected receive subsequent treatment elsewhere for the complaint which prompted the initial CAMHS referral. An audit of rejected referrals in a number of Scottish NHS health boards found that a large majority of rejected referrals were classed as ‘unsuitable’ (i.e. they did not meet the health board criteria for referral due to severity or the nature of the complaint) or that not enough information was supplied in the referral for it to proceed (26).

### How does it compare with other work?

#### Sex/Gender

Epidemiological studies of neurodevelopmental and mental health disorders and service use in children and young people consistently report gender differences. This study is concordant with national reporting for the whole population (including adults) which found markedly higher levels of mental health prescribing for females than males, apart from medicines used to treat ADHD (16). Open data for CAMHS referrals do not include a breakdown by sex so it is not possible to compare the pattern of increasing referrals for females in Grampian following the pandemic with the national picture. However, an audit of rejected referrals in seven Scottish health boards found that males accounted for a larger proportion (54%) than females, which is concordant with the findings of this work. The audit was conducted over a single year, so it is not possible to comment on trends through time.

A rapid literature review conducted in 2019 by the Scottish Government (23) looked specifically at evidence of worsening mental wellbeing among adolescent girls in Scotland. This found evidence of generally increasing prevalence of self-reported poor mental wellbeing in a variety of indicators among adolescents generally, but particularly for adolescent girls. The review concluded that there is little robust causal evidence to explain this observation.

There are competing hypotheses about whether the observed differences in mental health service use reflect *natural* differences in incidence and prevalence of mental health disorders between boys and girls, socially constructed expectations around mental health and behaviours, or the way these expectations influence professionals who make or assess referrals. Boys and girls are thought to manifest their mental health, social and behavioural patterns in different ways (37), although it is unclear the extent to which this is a consequence of biological or social processes. There is also concern that neurodevelopmental diagnostic criteria may not adequately reflect symptom manifestations in females (38), which may lead to delays in or lack of diagnosis and/or treatment compared with younger boys (39).

### Prevalence of poor mental health

Alongside our findings of increasing mental health prescribing and CAMHS referrals in children and young people over time, other work has observed a national increase in the prevalence of poor mental health (40). The Scottish Schools Adolescent Lifestyle and Substance Use Survey (SALSUS), which looks specifically at 13- and 15-year olds found continuous rises in the proportion with borderline or abnormal responses to the Strengths and Difficulties Questionnaire (SDQ) from 2010 to 2018 (41). They also found that the average Warwick-Edinburgh Mental Wellbeing Scale (WEMWBS) had decreased between 2015 to 2018, which suggests worsening mental wellbeing. Both indicators also had clear patterns by sex, with 15-year-old girls having the highest rate of borderline/abnormal SDQ and the lowest average WEMWBS scores. The Scottish Health Survey (SHeS) also looked at WEMWBS scores in children aged between 13 and 15 and found higher mental wellbeing scores in boys than girls (7). SALSUS reported social gradients in both SDQ and WEMWBS scores by deprivation, with the highest rates of poor mental health being found in the most deprived areas.

### Potential Mechanisms

The aims of this study are descriptive in nature and it is not possible to determine the mechanisms which produce the observed trends. For instance, increasing mental health prescribing and referrals to CAMHS could result from increasing prevalence of mental health disorders in the population, improved recognition of the need to seek help among young people and their carers, or improved access to treatments and referral pathways.

The same is also true of sex differences in referral and treatment for mental health conditions, although increasing CAMHS referral in girls but not boys since the start of the COVID-19 pandemic should prompt further investigation. Patterns of help-seeking behaviour may differ between boys and girls and this may also interact with age. Girls tend to appear in these records at older ages and in larger numbers, whilst boys are more likely to be in younger age groups. It may be that girls present later because their needs are not recognised at earlier ages, or that boys in need are less likely to be referred for or receive treatment once they reach adolescence.

Local changes in the demographics of the population treated by CAMHS, with higher rejections for younger boys and higher acceptances for older girls, predated the onset of the COVID-19 pandemic. Given the roughly static number of accepted referrals throughout the study period, this change in those accepted for treatment may reflect a conscious change in service prioritisation or greater levels of clinical acuity in specific population groups. Without more detailed information around reason for referral or rejection, this is not possible to determine.

Higher incidence of mental health prescribing and referral for specialist CAMHS for children in the most deprived areas has been persistent throughout the study period. The mechanisms through which area-level deprivation could influence mental health are varied (42) and include a lack of access to material resources, support services and socioeconomic opportunities which might improve mental wellbeing.

### Unanswered Questions and future research

Future work should explore the incidence and prevalence of diagnosable mental health conditions to allow for comparison with patterns of increasing mental health prescription and increasing demand for specialist CAMHS. This would help to determine whether patterns from administrative and service use sources reflect underlying trends in incidence or changes in recognition of mental health conditions and help-seeking behaviour.

More comprehensive data from primary care and other mental health support services available to children and young people should be made available for future research. In particular, community-based services (e.g. CAMHS tier 1 and 2) and GP data should be prioritised as these are likely to be the first point of contact with health professionals for people in need of support around mental health. Data from educational settings and charitable organisations providing mental health support are also likely to provide useful information. Future analysis of mental health prescribing and CAMHS referral should incorporate more detailed information related to clinical indications as well as reasons for referral and rejection. Public involvement in this work highlighted that deeper analysis of rejected referrals in the future was important and that this should explore patient pathways using routine sources of data as well as make use of qualitative methods to explore help seeking behaviours in children and their carers.

Whilst we have been able to describe patterns in prescribing and CAMHS referral by some demographic and socioeconomic characteristics, other important factors have not been possible to investigate. The limited availability and poor quality of ethnicity indicators in administrative healthcare records may underestimate inequalities (43) and individual or household indicators of socioeconomic position were not available to us. Future research should look to link reliable sources of social data with administrative health records to better understand the complex mechanisms which influence child and adolescent mental health and service/treatment access.

## Conclusions

The findings of this work indicate there is a bottleneck between rising mental healthcare need at the primary level (prescriptions) and the static size of the population of children treated by specialists in tertiary care. This study also found substantial differences in care use between the sexes across age groups, and much higher need in the most socioeconomically deprived communities. Children’s mental health care services should consider structural changes in provision which account for different patterns of need for these different populations. Frequent monitoring of care use is needed, as indicated by the rapid changes we found in specialist CAMHS referrals for older adolescent girls since the start of the COVID-19 pandemic.

## Supporting information

GRIPP2-SF

Credit Taxonomy

## Data Availability

All analysis was carried out in the Grampian Data Safe Haven (project ID: DaSH477) on pseudonymised individual-level data. Per Scottish Safe Haven Charter, only aggregate data can be released from the Grampian Data Safe Haven for publication, but all individual-level data will be archived for 5 years on project completion and may be accessed by application to the Grampian Data Safe Haven (email dash{at}abdn.ac.uk) on condition that appropriate project approvals were secured. Following the archiving period this data will be deleted.
Data analysis and figure generation was conducted using R (version 4.0.3) in RStudio (version 1.4.1103). All relevant analysis code files and summary data are available in the supplementary materials and are hosted on the project GitHub repository (https://github.com/AbdnCHDS/NDL_prescribing_referrals_paper).

## List of abbreviations

CAMHS: Child and adolescent mental health services
PIS: Prescribing Information System
SIMD: Scottish Index of Multiple Deprivation
DaSH: Grampian Data Safe Haven
TRE: Trusted Research Environment
CHI: Community Health Index
BNF: British National Formulary
SII: Slope Index of Inequality
RII: Relative Index of Inequality
ADHD: Attention Deficit Hyperactivity Disorder

## Declarations

### Ethics approval and consent to participate

This project was approved by the North Node Privacy Advisory Committee (NNPAC) (project ID: 6-063-21). NNPAC provides researchers with streamlined access to NHS Grampian data for research purposes and committee approval incorporates approvals from: the project sponsor, institutional ethics committee, the local Caldicott Guardian, and NHS Research & Development. It was not practicable to obtain informed consent from individual patients for this research which uses electronic patient records. Use of this pseudonymised unconsented patient information received ethical approval from the North Node Privacy Advisory Committee as detailed above. This research was conducted in a safe haven which adheres to the principles and standards outlined in the Charter of Safe Havens in Scotland (44) to ensure patient identity and privacy was protected. All research methods using human data were performed in accordance with the Declaration of Helsinki

### Consent for publication

Not applicable.

### Availability of data and materials

All analysis was carried out in the Grampian Data Safe Haven (project ID: DaSH477) on pseudonymised individual-level data. As per Scottish Safe Haven Charter, only aggregate data can be released from the Grampian Data Safe Haven for publication, but all individual-level data will be archived for 5 years on project completion and may be accessed by application to the Grampian Data Safe Haven (email dash{at}abdn.ac.uk) on condition that appropriate project approvals were secured. Following the archiving period this data will be deleted.

Data analysis and figure generation was conducted using R (version 4.0.3) in RStudio (version 1.4.1103). The datasets supporting the conclusions of this article are available in the project GitHub repository (https://github.com/AbdnCHDS/NDL_prescribing_referrals_paper or https://doi.org/10.5281/zenodo.6655991).

### Authors Contributions

Contributor Roles Taxonomy (CRediT) (45) has been used to make author contributions explicit and is available in the supplementary materials (Table S2) accompanying this publication. All authors (WB, CB, SG, BO, SP, AR, DR, HR, MR, ET, KW & JB) were involved in the conceptualisation as well as review and editing of this article. WB wrote the original draft. WB and JB conducted the formal analysis and investigation, wrote software, performed validation and produced visualisations. Data curation was performed by HR, WB, JB and KW (on behalf of Grampian DaSH). Funding was acquired by JB, CB, SG and KW. Methodology was developed by WB, JB, CB and SP. Project administration was conducted by JB, CB, SP and KW. Resources were provided by DR, ET, KW and HR. Supervision was performed by JB, CB and SP. Patient and public engagement activities were co-ordinated by MR and SG and BO (on behalf of the ACHDS PPIE group), WB and JB took part in these activities.

## Acknowledgements

We thank The Health Foundation for providing financial support to allow this analysis to be conducted and for facilitating the Networked Data Lab partnerships which have informed this work. The Health Foundation is an independent charity committed to bringing about better health and healthcare for people in the UK. We thank the NHS Grampian Health Intelligence team for facilitating use of this data. We also thank the Grampian Data Safe Haven (DaSH) for processing the data used in this study, as well as providing the secure platform used in this analysis and administrative support. We are grateful to members of the public who have made invaluable contributions to this work. This work uses data provided by patients and collected by the NHS as part of their care and support. We thank the anonymous reviewers for their careful reading of this manuscript and for their constructive comments.

## Public Involvement

There is widespread agreement that public participation is necessary for health data-intensive projects (46). We formed, trained, and involved a PPIE group of nine persons interested in health data usage for public benefit through a series of interactions throughout the research cycle including narrowing the focus of the research topic, developing analytical plans, sense-checking interpretations and improving the readability and accessibility of the results.

We also held group interaction with nine carers/parents and community workers who work with families and children. They reviewed near-final findings, remarked on usefulness and relevance, and suggested areas for further investigation. A comprehensive report of this effort utilising the GRIPP-2 reporting checklist for public involvement (47) is available in the supplementary materials (Table S1).

