## supplementary table S3 for "Inequalities in children’s mental health care: analysis of routinely collected data on prescribing and referrals to secondary care"

Table S3. List of mental health medications by British National Formulary Section and Subsection

| BNF Section Code | Section Name | BNF Subsection Code | Subsection Name | Approved Name |
|---|---|---|---|---|
| 401 | **Hypnotics and Anxiolytics** | 40101 | ***Hypnotics*** | Melatonin |
| | | | | Zopiclone |
| | | | | Temazepam |
| | | | | Nitrazepam |
| | | | | Zolpidem |
| | | | | Chloral Hydrate |
| | | | | Cloral Betaine |
| | | | | Sodium Oxybate |
| | | 40102 | ***Anxiolytics*** | Diazepam |
| | | | | Lorazepam |
| | | | | Buspirone Hydrochloride |
| 402 | **Drugs used in psychoses and related disorders** | 40201 | ***Antipsychotic drugs*** | Olanzapine |
| | | | | Risperidone |
| | | | | Aripiprazole |
| | | | | Chlorpromazine Hydrochloride |
| | | | | Quetiapine |
| | | | | Haloperidol |
| | | | | Sulpiride |
| | | | | Levomepromazine |
| | | | | Trifluoperazine |
| | | | | Promazine Hydrochloride |
| | | | | Lurasidone Hydrochloride |
| | | 40202 | ***Antipsychotic depot injections*** | Aripiprazole |
| | | 40203 | ***Drugs used for mania and hypomania*** | Lithium Carbonate |
| | | | | Sodium Valproate |
| 403 | **Antidepressant drugs** | 40301 | ***Tricyclic and related antidepressant drugs*** | Amitriptyline |
| | | | | Nortriptyline |
| | | | | Clomipramine Hydrochloride |
| | | | | Imipramine Hydrochloride |
| | | | | Lofepramine |
| | | | | Trazodone Hydrochloride |
| | | | | Dosulepin Hydrochloride |
| | | 40302 | ***Monoamine-oxidase inhibitors*** | Phenelzine |
| | | 40303 | ***Selective serotonin re-uptake inhibitors*** | Sertraline |
| | | | | Fluoxetine |
| | | | | Citalopram |
| | | | | Escitalopram |
| | | | | Fluvoxamine Maleate |
| | | | | Paroxetine |
| | | 40304 | ***Other antidepressant drugs*** | Mirtazapine |
| | | | | Duloxetine |
| | | | | Flupentixol |
| | | | | Venlafaxine |
| | | | | Agomelatine |
| | | | | Vortioxetine Hydrobromide |
| 404 | **CNS Stimulants and drugs used for ADHD** | 40400 | ***CNS Stimulants and drugs used for ADHD*** | Methylphenidate Hydrochloride |
| | | | | Atomoxetine |
| | | | | Lisdexamfetamine Dimesylate |
| | | | | Dexamfetamine Sulfate |
| | | | | Modafinil |
| | | | | Guanfacine Hydrochloride |
| 410 | **Drugs used in substance dependence** | 41001 | ***Alcohol dependence*** | Acamprosate Calcium |
| | | 41002 | ***Nicotine dependence*** | Nicotine |
| | | | | Varenicline Tartrate |
| | | 41003 | ***Opioid dependence*** | Methadone Hydrochloride |
| | | | | Buprenorphine And Naloxone |
