## Supplementary material for "Inequalities in children’s mental health care: analysis of routinely collected data on prescribing and referrals to secondary care": GRIPP2-SF

**Table S1**
Patient and public involvement (PPI) in this study, described using the GRIPP 2-SF checklist (Staniszewska et al., 2017)

| Section and topic | Item |
|---|---|
| **1: Aim**<br>Report the aim of the study | We wanted to look at the mental health care offered for children and young people in the Grampian Health Board region using healthcare administrative information on community prescriptions and referrals to outpatient Child and Adolescent Mental Health Services (CAMHS) (from 2015 to 2021). We ensured this information would be accessed safely without seeing names, dates of birth or addresses. We set out to study the types of care young people and children received, to see if there are disparities in who received care and to compare care before and after the COVID-19 pandemic. |
| **2: Methods**<br>Provide a clear description of the methods used for PPI in the study | Our PPI leads co-developed a PPI framework based on the NIHR: National Institute for Health and Care Research guidance with the funder, and a local plan of specific PPI activities. We created and trained a patient and public involvement and engagement group (known as the ACHDS PPIE group), including nine people from the local community, with diverse backgrounds and experiences (i.e., a neuroscience and psychology student, retired science teacher, retired accountant, retired chemist, retired engineer, retired schoolteacher, cancer nurse, volunteers supporting young people and people with mental health problems, carer, cancer survivor, a patient with a chronic condition). All are interested in promoting health data use for health and social care improvement. We interacted with the group through online group discussions and via emails. We consulted the group on the planned activities. The group assisted the research team at all project development and execution stages. They were involved in refining the focus of the research questions, developing a plan of analysis, and sense-checking interpretations of results. They checked outputs for readability and comprehension. Members of the group contributed to edits of articles and are co-authors. The group made suggestions of contacts in their networks who may have relevant insights or experience to contribute to the project and on how to share the findings with a broader public. Following the group's advice, we hosted an online discussion with people who have relevant experience in supporting young people – we called 'advocates of young people' (including carers or parents and people working with families and children in the community). This group of advocates for young people was involved in checking if the near-final findings make sense, if they are useful and relevant. Both groups made suggestions for further research. |
| **3: Results**<br>Outcomes—Report the results of PPI in the study, including both positive and negative outcomes | <u>When the analytical team presented the initial research question to the ACHDS PPIE group followed by a group discussion, the ACHDS PPIE group:</u><br>• Highlighted the importance of understanding the *"context"*: what services are available (e.g., rural vs urban differences in access), how easily services could be accessed, *"who are we missing out there"* because of unavailable or incomplete data.<br>• Suggested to link data, if possible, to provide a more complete picture, for example, looking at school records.<br>• Recommended engaging with young people and those who surround them for sense checking our interpretations (i.e., "*making sure we do not misrepresent the stories of real people behind the numbers*").<br><u>When presented with the interim results for mental health prescribing data during a group discussion, the ACHDS PPIE group provided the following:</u><br>*Feedback on methods of presenting the findings:* |

| Section and topic | Item |
|---|---|
| | • The concept of rate that we were concerned the public may not understand, the group found understandable, regardless of the person's background, because rates have been frequently used in communication about COVID-19 infections.<br>• We were told our ability to describe findings using plain English has improved.<br>• When the SIMD (Scottish Index of Multiple Deprivation) was introduced, they suggested we explain the acronym SIMD; we use the term 'factors' instead of 'domains' (which the public will be less familiar with) and add labels to the legend.<br>• The group liked graphs to show trends over time.<br>• Suggested that tables are too complex for the public to understand, suggested that each table would need to be explained separately and offered ideas about what could be discussed.<br><br>*Commented on unexpected findings:*<br><br>• *"How come people in deprived areas were prescribed more given the known issue with access to services - one of the SIMD factors?"* – the group suggested the need to highlight that prescription is the most basic and most readily available treatment, and most of those prescriptions would be through a general practitioner. The group felt it would be good to provide, is possible, the source of prescribing.<br>• *"Those 26 MH prescriptions per 100 people in less deprived areas still sounds like a lot"* - the group suggested we consider if the findings are comparable to the national average and overall acknowledge the limitations making it clear that this work is about the extent of the problem and does not cover why medications were prescribed and whether prescribing was beneficial.<br><br>*Suggested considerations for interpretations:*<br><br>• *"What's the definition of prescription?"* – state it upfront and consider the benefits (such as good completion) and downsides (such as referral source data is unavailable) of that administrative data used.<br>• *"What about other mental health treatments?"* – consider that there are other forms of treatment (such as talking therapy) and that information is limited or not included in the analysis.<br>• *"How come ADHD is included in mental health disorders analysis?"* - the term 'mental health prescriptions' can be misleading, so state what classification is used.<br>• "*What about age and sex differences*?" - A graph showing a type of prescription and age would be useful and consider looking at the impact of the length of prescribing and new versus reoccurring prescriptions.<br>• *"What's the effect of parental influence?"* - while children can refuse treatment from the age of 12, they can be expected to be under the influence of parents.<br><br>*Advised whom we should speak to next and how*:<br><br>• The group advised to speak to adults surrounding young people: support groups (such as charities), schools, and non-statuary organisations (such as sports groups), including those operating in more deprived areas. The group advised we should ask those people: What has surprised you? Does it echo your experiences? What could be done about it? Based on the types of queries this group had, it |

| Section and topic | Item |
|---|---|
| | would also be important to explore with people surrounding young people, what aspects of those findings we should try to understand better (to make recommendations for future research).<br>• Advised to consider speaking to young people when results are finalised.<br>• Advised that young people probably do not need anything beyond plain English statements; those supporting them would appreciate the map and simple graph (less so the table).<br><br><u>Summary from a group discussion about the near-final results (including prescribing and referral data) with nine 'advocates of young people':</u><br><br>The results presented echo their experiences. They were surprised with rejection rates of referrals to CAMHS; but contrary to what we thought, while the rejection rates increased, they expected these to be even higher. They observed that mental health needs increase in girls at around the age of 13 overlap with other patterns of behaviour at that age (like older girls losing interest in sports and experimenting with drugs and alcohol). Sex and deprivation differences, they said, need to be interpreted carefully, given the numbers may reflect differences in opportunities and abilities in access to care (such as implicit bias, health literacy, help-seeking behaviour, and emotional literacy), which are related to underlying complex socioeconomic and psychological factors (such as family size, mental health history of parents). Early prevention interventions at a school level, such as free-of-charge activities to promote good mental health for children of all ages, would be helpful. Also, the message was that "*children must be seen earlier*" and a systemic approach is needed (i.e., an organisation/system/policy-level intervention), through targeting key people surrounding young people (like investment in non-expert staff's mental health support skills, advice for families about care access and finances, improving awareness of healthcare professionals, opening referrals for teachers). It is evident that COVID-19 has had a negative impact on mental health and/or care locally, and this information was needed to inform resource allocation planning.<br>The results were thought to raise a series of research questions, such as: What are the contextual influences on onset of mental health issues and help seeking? Are diagnostic or referral criteria, or professional or implicit bias somehow contributing to gender bias in diagnosis of depression, ADHD, and eating disorders? How do we do support children to make sure they do not need referrals or are supported appropriately when referrals are rejected? What are the transition patterns between universal services (schools, GP, health visiting, outpatient care and so on) and quality of support they offer?<br><br><u>When the near-final results (including prescribing and referral data and the summary from the discussion with 'advocates of young people') were discussed with the ACHDS PPIE group, the group provided:</u><br><br>*Feedback on methods of presenting for the public*<br><br>• To change the term 'secondary care'- it is unclear, and the word secondary is also used in the context of education<br>• The group preferred the graphs over the tables they saw last time - they liked the selected graphs, especially trend over time charts but grouped bar charts were felt to be hard to read for a person with dyslexia and suggested to try back-to-back bar graphs.<br>• For the reporting and dissemination purpose, graphics need to be seen clearly, so they need to be in higher resolution with bigger font and one chart per slide. |

| Section and topic | Item |
|---|---|
| | • They liked how first a key finding was stated in simple terms (e.g., 'Boys receive more mental health prescriptions in younger ages and girls receive more in older ages')<br>• They said the graphs used are too alike, so a mixture of graphs and infographics would be welcome.<br><br>*Shared general impressions of findings:*<br>• The group described the findings as *"interesting", "disturbing"*, *"amazing"*, *"blown away by the figures"* (especially age and sex differences).<br><br>*Made considerations for interpretations:*<br>• This data must be contextualised, so it is essential to compare it with national rates or other regions.<br>• Can we compare children in the city and urban areas?<br>• Why did rejection rates increase (e.g., not suitable, a matter of capacity, eligibility)? The same person can be referred several times before they get seen, is this captured and accounted for?<br><br>*The group felt that the findings "raise more questions than they answer…and more research is important", and suggested the following questions warrant further research:*<br>• How will those experiences affect young people in the future? If more girls are seen in specialist care while boys hide problems, don't address, and don't ask or get help – what's the impact?<br>• Important to understand why young people are referred/rejected?<br>• What is the impact and relevance of a lack of school provision during the pandemic on the observed trends?<br>• What role can school play in addressing the observed problem? Investment in schools was a recurring theme.<br>• Increasing numbers of children with ADHD in primary school (Do girls with ADHD present differently? What happens to young people with ADHD after school age – "*ADHD doesn't go away*"?)<br>• What is the potential role of experimenting with alcohol and drugs on those changes?<br>• What is the mechanism for delivering changes? What upskilling is required?<br>• What is the potential role of a decrease in interest in the sport in older girls on the observed effects?<br>• Groups we should focus on (What about children that are missed by the system? What about vulnerable children?)<br><br>Throughout the write-up phase for both the results paper and the methods paper, the ACHDS PPIE partners contributed to the lay sections and contributed to edits of the briefing paper and this article. |
| **4: Discussion**<br>Outcomes—Comment on the extent to which PPI influenced the study | Public contributors' advice resulted in additional analysis (such as stratifications by sex and age) and methodological considerations made (for a type of prescription, lengths of receiving medication, and new versus repeat referral). People who surround young people sense-checked and contextualised our interpretations and drew implications for research. |

| Section and topic | Item |
|---|---|
| overall. Describe positive and negative effects | The ACHDS PPIE group also contributed significantly to improving the readability and accessibility of outputs, which each time served as an opportunity to reflect on the meaning of our work. They helped us develop lay-person-friendly ways of presenting data, reviewed outputs (i.e., presentations, lay summaries for project protocol and permissions, conference abstract, social media posts, project reports), and co-designed plans for involvement and engagement with target groups of public members.<br><br>Overall, public involvement in this study influenced essential aspects of the study and the following stages of our research cycle. This might have been related to several factors: 1) we have engaged with best practice guidance to decide what PPI will be done and how, 2) we have trained our ACHDS PPIE group, providing them with a 'language' needed to talk about data science and PPI), 3) ACHDS PPIE group was involved from the beginning which helps to create a sense of shared ownership and meaning, 4) we have taken detailed notes on and summarised all of our conversations, as well as produced actionable points from each interaction, proving that their contributions are valued and relevant. The right collective skills of PPI leads were essential, such as ample experience in facilitating group discussions, conducting text analysis, communicating clearly and with a purpose in a friendly and accessible manner, and experience in interdisciplinary collaborations. Having the right context, i.e., support from other PPI leads across the Networked Data Lab (NDL) programme, funding to finance public contributors' time, and a supportive attitude of their involvement from the analytic team also assisted in the positive impact that PPI had on this study. Due to a lack of good practise examples, we had to create our PPI workstream from the ground up and at a rapid pace to meet funder deadlines, which was difficult and necessitated a new method of working within the team to enable an effective multi-disciplinary approach. The buy-in of our analytical team, as well as a noticeable shift in their appreciation of and for PPI, is a definite advantage. We have yet to implement our ACHDS PPIE group's recommendation to diversify the group's demographics by involving people from underrepresented communities (hard to reach groups), and we have not included young people with lived experience (for safeguarding reasons), so we have missed out on some critical perspectives. |
| **5: Reflections**<br>Critical perspective—Comment critically on the study, reflecting on the things that went well and those that did not, so others can learn from this experience | In our experience PPI is more effective with a whole system approach deploying established methods, such as those used in social sciences. Moreover, we observed the complexity and importance of the communication underpinning PPI and will develop this as a research theme. Developing a PPI framework using robust methods and building an effective public involvement communication model took significantly more time than initially anticipated. At this point, we find the set-up that works best for us is with a PPIE Lead responsible for critical thinking, planning, interpretation, and reporting (half day a week), assisted with management (communications, group discussions, actionable summaries from interactions - one and a half days per week) and administrative (emails, diary invites, PPI partner reimbursement - one and a half-day per week) support. As we continue to develop the PPI workstream within the NDL programme, where appropriate, we will involve people with lived experience earlier in the process. |
