## Supplementary material for "Inequalities in children’s mental health care: analysis of routinely collected data on prescribing and referrals to secondary care": Credit Taxonomy

Table S2. Credit taxonomy for author contributions.

| Term | Description | Authors |
|---|---|---|
| Conceptualization | https://credit.niso.org/contributor-roles/conceptualization/ | All authors |
| Data curation | https://credit.niso.org/contributor-roles/data-curation/ | HR, WB, JB, KW on behalf of Grampian DaSH |
| Formal analysis | https://credit.niso.org/contributor-roles/formal-analysis/ | WB, JB |
| Funding acquisition | https://credit.niso.org/contributor-roles/funding-acquisition/ | JB, CB, SG, KW |
| Investigation | https://credit.niso.org/contributor-roles/investigation/ | WB, JB |
| Methodology | https://credit.niso.org/contributor-roles/methodology/ | WB, JB, CB, SP, |
| Project administration | https://credit.niso.org/contributor-roles/project-administration/ | JB, CB, SP, KW |
| Resources | https://credit.niso.org/contributor-roles/resources/ | DR, ET, KW, HR |
| Software | https://credit.niso.org/contributor-roles/software/ | WB, JB |
| Supervision | https://credit.niso.org/contributor-roles/supervision/ | JB, CB, SP |
| Validation | https://credit.niso.org/contributor-roles/validation/ | JB, WB |
| Visualization | https://credit.niso.org/contributor-roles/visualization/ | WB, JB |
| Writing – original draft | https://credit.niso.org/contributor-roles/writing-original-draft/ | WB |
| Writing – review & editing | https://credit.niso.org/contributor-roles/writing-review-editing/ | All authors |
| Patient and Public Engagement | | MR, SG, BO, WB, JB |
