## Supplementary figures and images for "Inequalities in children’s mental health care: analysis of routinely collected data on prescribing and referrals to secondary care"

### supplementary figure S1

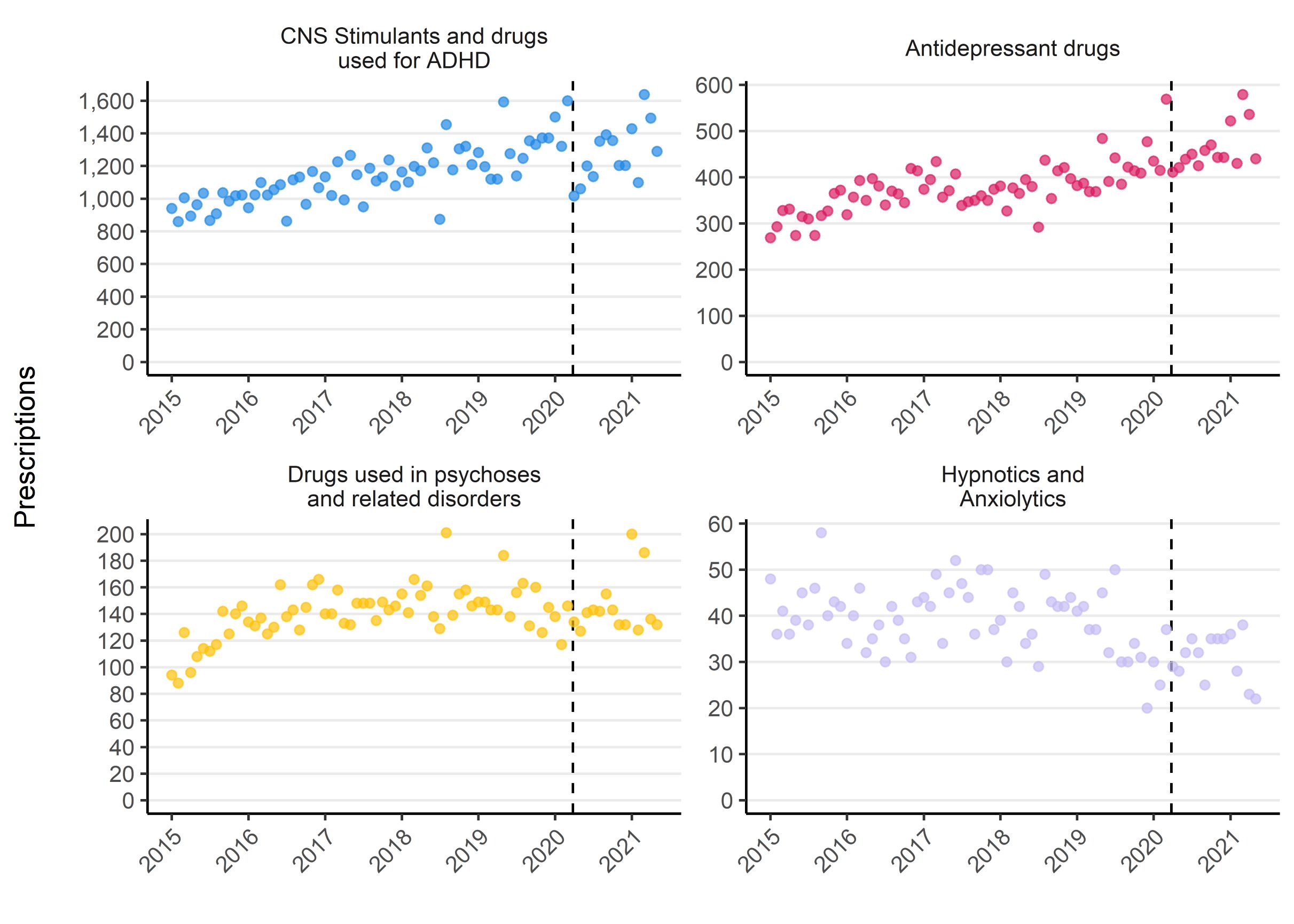

### supplementary figure S2

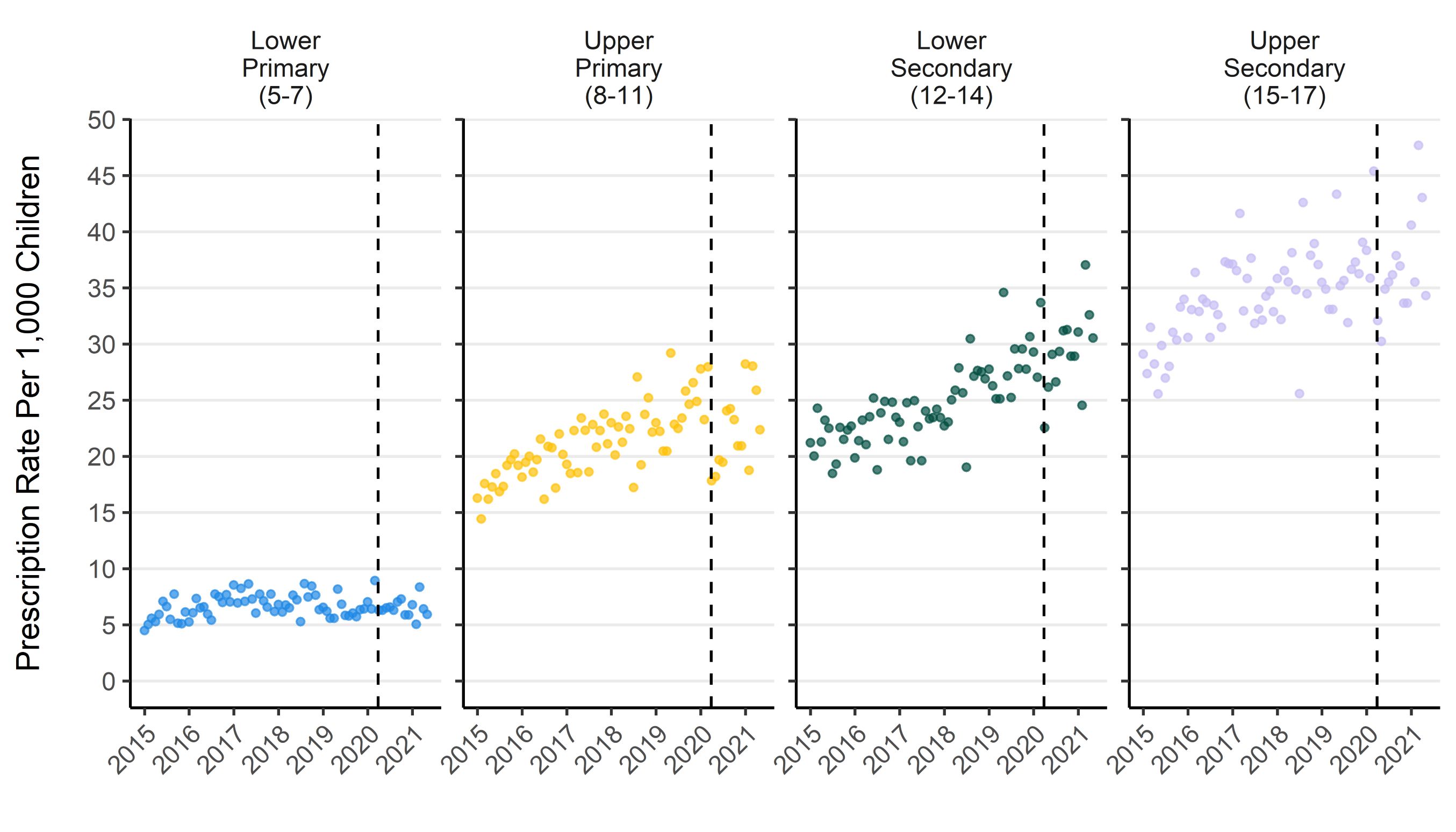

### supplementary figure S3

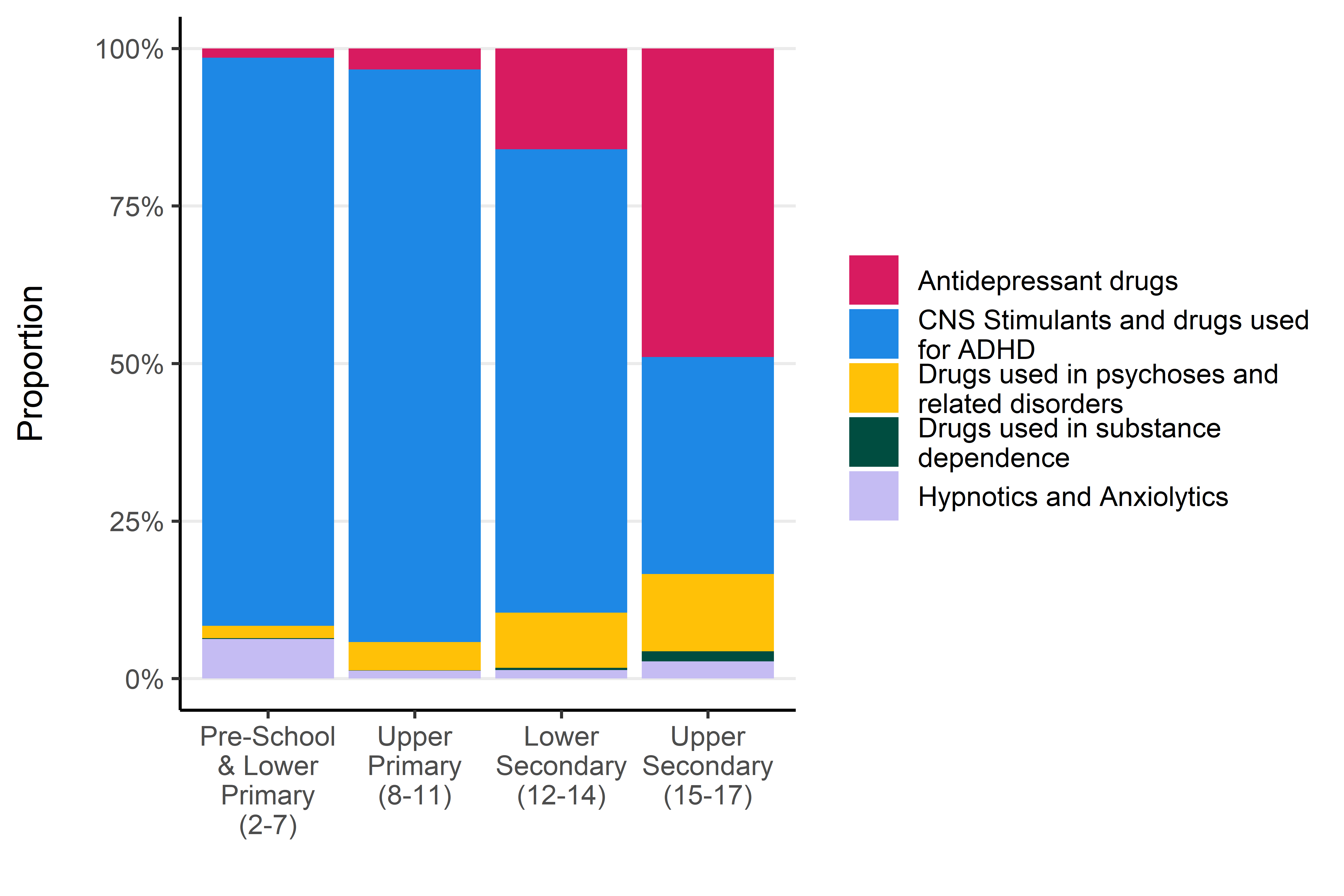

### supplementary figure S4

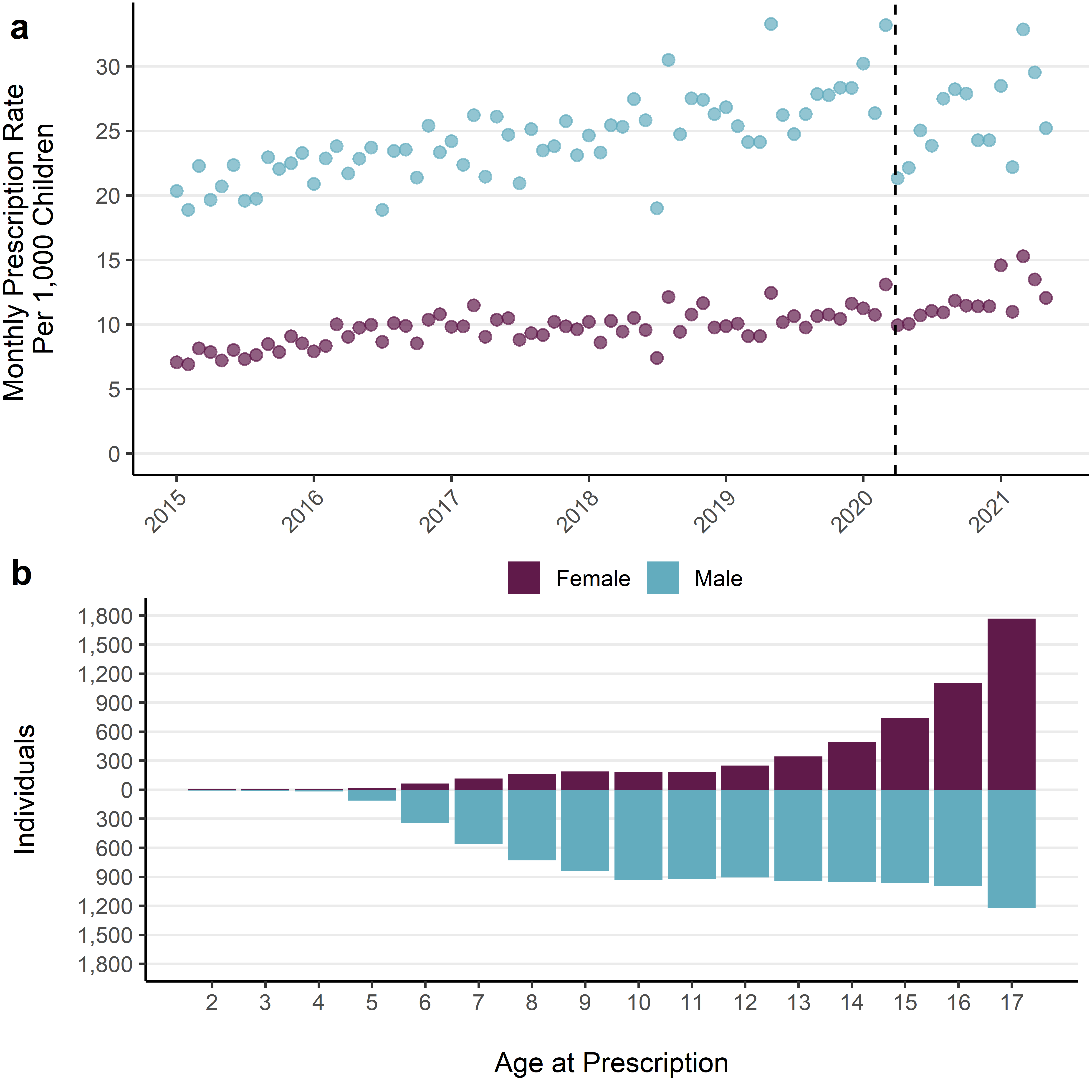

### supplementary figure S5

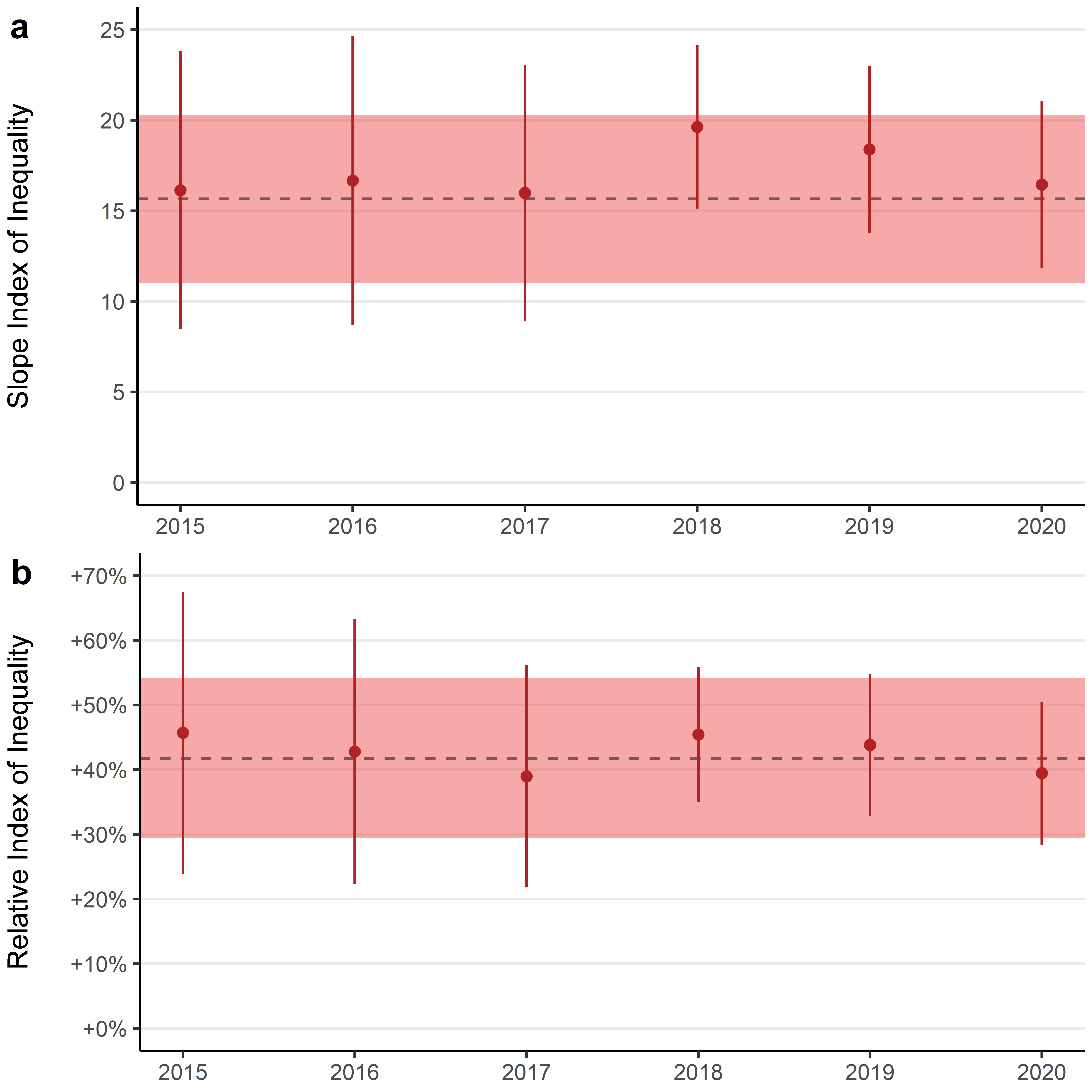
